# Estimating the cost-effectiveness of maternal vaccination and monoclonal antibodies for respiratory syncytial virus in Kenya and South Africa

**DOI:** 10.1101/2022.09.09.22279780

**Authors:** Mihaly Koltai, Jocelyn Moyes, Bryan Nyawanda, Joyce Nyiro, Patrick Munywoki, Stefano Tempia, Xiao Li, Marina Antillon, Joke Bilcke, Stefan Flasche, Philippe Beutels, D. James Nokes, Cheryl Cohen, Mark Jit

**Affiliations:** Department for Infectious Disease Epidemiology, London School of Hygiene & Tropical Medicine, London, UK; Centre for Mathematical Modelling of Infectious Diseases, London School of Hygiene & Tropical Medicine, London, UK; Kenya Medical Research Institute (KEMRI) - Center for Global Health Research, Kisumu, Kenya; Division of Global Health Protection, Centers for Disease Control and Prevention, Nairobi, Kenya; KEMRI-Wellcome Trust Research Programme, KEMRI Centre for Geographical Medicine Research-Coast, Kilifi, Kenya; Centre for Respiratory Disease and Meningitis, National Institute for Communicable Diseases, Johannesburg, South Africa; School of Public Health, Faculty of Health Sciences, University of the Witwatersrand, Johannesburg, South Africa; Centre for Health Economic Research and Modelling Infectious Diseases (CHERMID), Vaccine & Infectious Disease Institute, University of Antwerp, Antwerp, Belgium; School of Life Sciences, University of Warwick, Coventry, UK

**Keywords:** respiratory syncytial virus, cost-effectiveness analysis, maternal vaccination, monoclonal antibodies, disease burden, hospital data, ARI, SARI

## Abstract

**Background:** Respiratory syncytial virus (RSV) causes a substantial burden of acute lower respiratory infection in children under 5 years, particularly in low- and middle-income countries (LMICs). Maternal vaccine (MV) and next-generation monoclonal antibody (mAb) candidates have been shown to reduce RSV disease in infants in phase III clinical trials. The cost-effectiveness of these biologics has been estimated using disease burden data from global meta-analyses, but these are sensitive to the detailed age breakdown of paediatric RSV disease, for which there have previously been limited data.

**Methods:** We use original hospital-based incidence data from South Africa and Kenya collected between 2010 and 2018 of RSV-associated acute respiratory infection (ARI), influenza-like illness (ILI), severe acute respiratory infection (SARI) as well as deaths with monthly age-stratification, supplemented with data on healthcare-seeking behaviour and costs to the healthcare system and households. We estimated the incremental cost per DALY averted (incremental cost-effectiveness ratio or ICER) of public health interventions by MV or mAb for a plausible range of prices (3-30 USD for vaccines and 6-60 USD for monoclonals), using an adapted version of a previously published health economic model (McMarcel) of RSV immunisation.

**Results:** Our data show higher disease incidence for infants younger than 6 months of age in the case of Kenya and South Africa than suggested by earlier projections from community incidence-based meta-analyses of LMIC data. Since MV and mAb provide protection for these youngest age groups, this leads to a substantially larger reduction of disease burden and therefore, more favourable cost-effectiveness of both interventions in both countries. Specifically, using published efficacy data, our mean estimate for reducing RSV-associated deaths in children under 5 years of age is 9% for MV and 28% for mAb in Kenya. In South Africa, the reduction is larger, with the mean estimate of 14% for MV and 48% for mAb.

In the case of the lowest dose prices (3 USD for MV and 6 USD for mAb), the healthcare system perspective ICERs per DALY averted drop to 144 USD (mAb) and 397 USD (MV) in Kenya, whereas it is net cost-saving from the perspective of the South African healthcare system. At the highest assumed dose prices of 30 USD for MV and 60 USD for mAb, the median estimates for the ICER are 4528 USD for MV and 2748 USD for mAb in Kenya, while in South Africa, it is 4694 USD for MV and 2566 USD for mAb.

**Conclusion:** Interventions against RSV disease may be more cost-effective than previously estimated following the incorporation of new data indicating that the disease burden is highly concentrated in the first 6 months of life in two African settings.

## Introduction

Respiratory syncytial virus (RSV) is a globally widespread [1], [2] endemic virus, which is the most common pathogen in children diagnosed with acute lower respiratory infections (ALRI) [3], [4]. Most symptomatic and severe cases are in children younger than 5 years, with infants being the most affected, as most severe cases occur in the first year of life. The disease burden is particularly high in low- and middle-income countries (LMICs), where the majority of global RSV attributable hospitalisations and deaths are concentrated [3], with latest estimates suggesting over 62% of RSV-associated hospitalisations and nearly all deaths globally in LICs and LMICs [47].

Both maternal vaccination (MV) and monoclonal antibodies (mAb) may confer protection against RSV disease in early life. There have been multiple efforts to develop a maternal vaccine [5]. The most advanced maternal vaccine candidate (by Novavax) showed protection against RSV LRTI-associated hospitalisation and severe hypoxemia [6]. While it showed significant protection against its primary clinical endpoint (medically significant LRTI up to 90 days of life) in South Africa, it did not meet its primary clinical endpoint across all study sites due to low efficacy figures observed in the arm of the trial conducted in the United States [7]. Consequently, its pathway to licensure and marketing is unclear. In late 2020 a new phase III trial of the maternal vaccine candidate GSK3888550A (GlaxoSmithKline) was launched [8], [9] but enrolment was subsequently stopped due to a safety signal [49], and another clinical trial by Pfizer was started in mid-2020 (trial running until 180 days of life) [10]. Positive topline results were announced in June 2021 of the phase III trial of the monoclonal antibody Nirsevimab (Sanofi), demonstrating protection against RSV disease in healthy infants [11], confirming previous results [12] that showed 70% and 78% efficacy against medically attended and hospitalised RSV-associated LRTI, respectively, in a 150-day trial of 1453 infants. Most recent (March 2022) published results from the Nirsevimab trial similarly showed an efficacy of 74.5% for medically attended RSV-associated LRTI [50]. Interim mean efficacy figures [13] for the Pfizer MV were above 80%, significantly higher than the previously published values for the Novavax MV.

With these preventive biologics against RSV infection showing promise and possibly becoming commercially available in the years to come, it is important to assess the cost-effectiveness of public health interventions via vaccines or monoclonal antibodies for infants. This is especially important in the context of LMICs which have the highest burden but also face financial constraints and competing demands on healthcare resources from other diseases. The only currently available prophylactic RSV treatment (palivizumab, a monoclonal antibody administered by monthly injections over the RSV season) is in use only in a few high-income countries where it is recommended exclusively to high-risk groups due to its high cost [14]. Because maternal and next generation (long half-life) monoclonal antibodies have a duration of protection of 3-6 months, the cost-effectiveness of both vaccines and monoclonal antibodies depends on the age distribution of RSV burden in infants. The monthly resolution of the incidence data of the current study and the separate estimates for all and severe RSV disease allow for a more detailed analysis than the larger age brackets used for infants in previous analyses [3].

In this study, we provide updated estimates for the cost-effectiveness of MV or mAb-based public health interventions to prevent RSV disease in children under 5 years of age in two African countries, one lower middle income (Kenya) and the other upper middle income (South Africa). A previous study by Li et al. [15] used estimates for RSV incidence in the community and in hospital from Shi et al. [3] to quantify the cost-effectiveness of these interventions in 72 Gavi-eligible countries, while accounting for uncertainty in parameters by using probabilistic sensitivity analysis. We are able to refine this analysis by using recently available finely (monthly) age-stratified estimates for RSV-associated respiratory infections, hospitalisations and deaths in Kenya and South Africa. We also use new data from the same sources on country-specific cost estimates for both countries.

## Methods

### Study population and data collection

The age-stratified incidence of RSV-associated non-severe (ARI) and severe (SARI) acute respiratory illness was estimated for both countries. The methodology for obtaining and calculating incidence is described in detail in the accompanying papers (Moyes et al. 2022, Nyawanda et al.l 2022) and briefly described below.

#### Kenya

To estimate the RSV-associated ARI burden in Kenya, methods previously used for estimating the burden of outpatient ILI and RSV-associated ILI in Western Kenya [16] were combined with those used in estimating the burden of SARI and influenza-associated SARI [17], [18].

ARI was defined as an acute (onset within 10 days before diagnosis) illness with cough, difficulty breathing, sore throat or runny nose. The definition for extended SARI, in line with the WHO’s RSV surveillance case definition, was hospitalisation with an acute respiratory infection (onset within 10 days) with cough or difficulty breathing. For non-hospitalised severe cases, ARI with pneumonia was used as a proxy for SARI. The definition for pneumonia was a cough and difficulty breathing for >2 days or a diagnosis of pneumonia by a health care worker (HCW) within the last year.

First, the overall ARI rate in Kenya was established to estimate RSV-associated ARI. To do this, data on healthcare-seeking behaviour from households reporting ARI in the Household Morbidity Surveillance (HMS) [19] were used to estimate the proportion of ARI cases seeking formal medical care, by collecting data from over 27 thousand participants at biweekly home visits and at Lwak Hospital (rural western Kenya) where free care was provided by dedicated study clinical staff, asking ill persons if and where they sought care. The total number of ARI cases per year were collected from households in the base area of Asembo (Siaya County, Nyanza Region) in the catchment area of St. Elizabeth Lwak Mission Hospital (LMH) and through the Population-Based Infectious Disease Surveillance system (PBIDS) [20]. In the latter study field workers visited households in rural western Kenya (Asembo) and an informal settlement in Nairobi every two weeks collecting recent illness information; participants (>50,000 persons) could access free high-quality care in a referral clinic at each site. The ARI rate in the base area was calculated by dividing the number of ARI cases visiting LMH in a year by the population of the surveillance catchment area, scaled by the proportion of ARI patients who seek care from HMS. This rate was projected to the larger base region (Nyanza) by the ratio of ARI cases seeking care in the region [21] to those seeking care in Asembo [19]. Risk factors for respiratory infections (such as air pollution, malnutrition, or HIV prevalence) [22] were used in a linear model to project the ARI rate to other regions, while also accounting for different regional levels of healthcare-seeking behaviour. Data on healthcare-seeking behaviour was from a Health Utilization Survey (HUS; unpublished data) conducted in Siaya County (Nyanza Region), Nakuru County (Rift Valley), Kakamega County (Western) and Marsabit County (Eastern) to estimate the proportion of ARI cases seeking care.

Next, virological testing in Kenya was performed at LMH and used to obtain the RSV-associated rates of ARIs. Nasopharyngeal and oropharyngeal (NPOP) samples were systematically tested for the genetic material of respiratory viruses by RT-PCR (polymerase chain reaction) in the base area (Asembo) for all patients meeting the ARI case definition during 2010-2014, thereafter aggregated to annual RSV positivity for scaling to missing years of testing for the base (Nyanza) region. Testing for ARIs was unavailable for other regions (Table 1), therefore the ratio of RSV-positivity in ARIs to SARIs in the base region was applied to the SARI testing data of other regions to get the RSV-positivity for ARIs in the other regions.

To estimate RSV-associated SARI in Kenya, first data were collected from children under 5 years of age visiting Kilifi County Referral Hospital (CRH), combined with residency data from participants in the Kilifi HDSS [23]. Similar to ARI, the rate of SARI in the base area was projected to other regions by including risk factors [22] in a linear model and using the estimates for healthcare-seeking behaviour. The proportion of individuals hospitalised with pneumonia from a health utilisation survey was used as a proxy for SARI hospitalisations. RSV-associated SARI was estimated from systematic testing of NPOP samples for RSV in the Coastal and Nyanza Regions. For other regions, some of the data were pooled from five hospitals.

Estimates for in and out-of-hospital RSV-associated deaths per 100,000 population for Kenya (Figure 3) were derived by applying Kenyan in-hospital CFR estimates and the ratio of in-hospital to out-of-hospital deaths in South Africa [24] to the in- and out-of-hospital rates of SARIs in Kenya (Figure 1), resulting in an approximately equal number of in- and out of hospital deaths (Nyawanda et al.2022).

**Figure 1.**
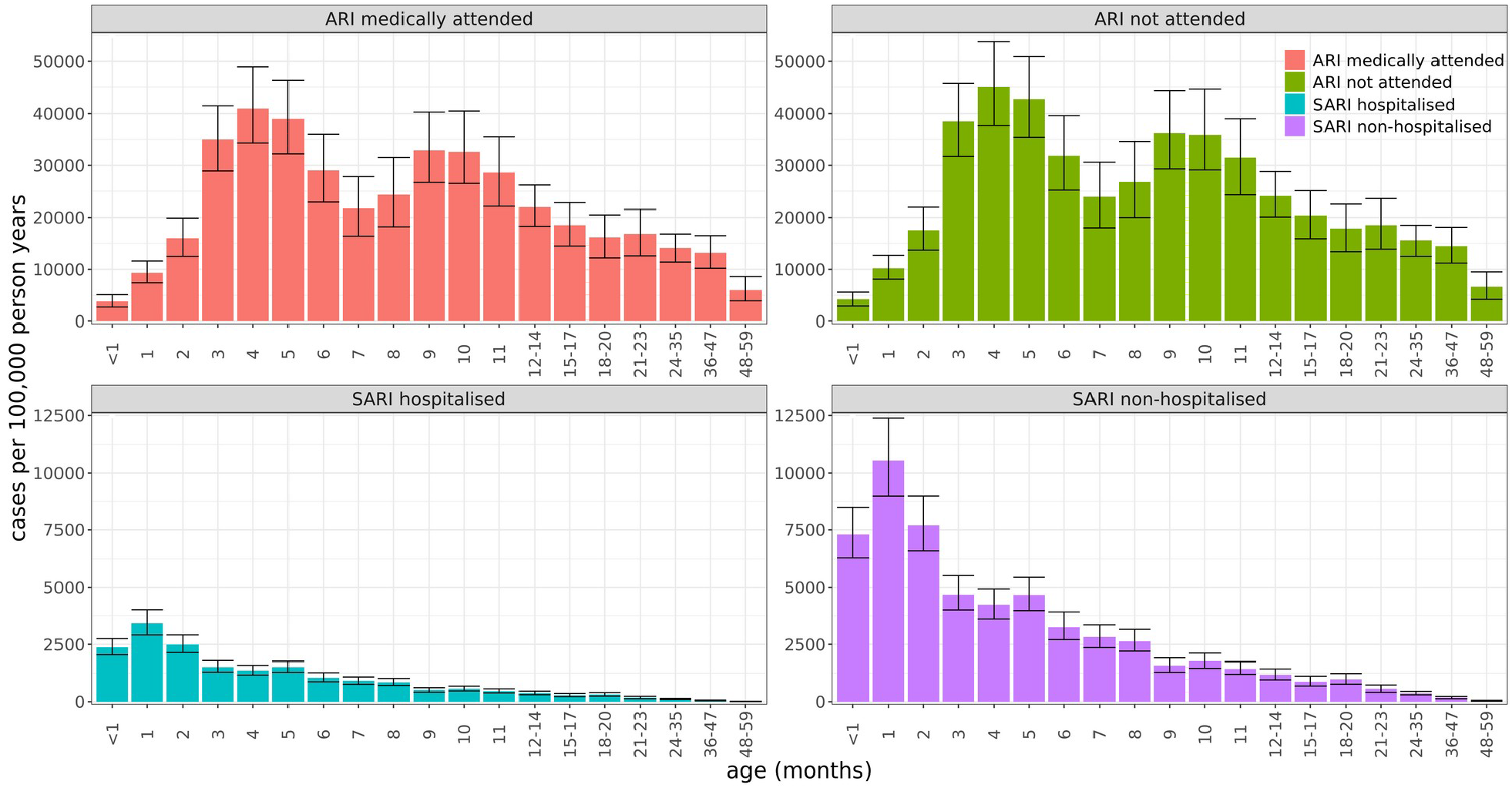
RSV-associated ARI and SARI rates per 100,000 person-years in Kenya by age group and medical status.

#### South Africa

The definition for hospitalised SARI was admission to hospital with a physician’s diagnosis of LRTI, with or without fever, and a duration of 10 days or less. For South Africa, there were two principal data sources, similar to the methodology of a previous study [25] on influenza. Surveillance was carried out in two public health outpatient clinics for ILI and in three public hospitals for ALRI in two provinces with a combined catchment area of nearly half a million people in 2015. Respiratory specimens were collected from all enrolled patients (ILI, SARI) and tested for 10 viruses, including RSV. RSV-positive co-infections were included. Secondly, healthcare utilisation surveys (HUS) [26], [27] were used for estimates of healthcare-seeking behaviour among individuals with reported ILI or SARI symptoms in the two communities. Similarly to Kenya, risk factors [17], [28], [29] (exposure to indoor air pollution, crowding, malnutrition, low birth-weight) were taken from the Demographic Health Survey (DHS) to project the SARI rates of the base region to other regions of the country.

For non-severe cases, RSV-associated influenza-like illness (ILI) cases were used. Using the same methodology as for SARI [25], cases were classified as ILI by case report forms for participants enrolled in the surveillance program and samples were transported to and RT-PCR-tested in the National Institute for Communicable Diseases (NICD) for 10 respiratory viruses, including RSV. The South Africa Demographic and Health Survey (DHS) [30] was used to account for risk factors to project local rates across provinces and calculate the national average. Age-specific rates of ILI were calculated from the corresponding SARI rates scaled by the proportion of SARI cases that sought outpatient care before hospitalisation and the ratio of hospital-referred to total ILI consultations [25]. ILI cases were defined as a person presenting with either temperature ≥38C or a history of fever and cough of a duration ≤10 days [31]. This is a narrower definition than the ARI definition in Kenya. To make the two case definitions comparable, we used data on the proportion of ARI cases with fever (23.3%), collected between 2015 and 2017 (2015: 33.3%, 2016: 20.5%, 2017: 16%) in Kenya [32], to broaden the ILI definition (by dividing by the proportion of ARI cases with fever) and align it with ARI. Figures in the main text are by using this broader definition.

Age-specific estimates of in-hospital and out-of-hospital RSV-associated deaths (Figure 3) were derived in a previous publication [24], also described in the accompanying paper by **Moyes et al. 2022**. In-hospital mortality was estimated by applying the observed case-fatality ratio to the national number of RSV-associated severe illnesses. Out-of-hospital mortality was estimated using published data on the proportion of deaths that occurred outside of hospital (26% in <5-year-old children).

### Calculation of disease burden, cost and cost-effectiveness

The cost-effectiveness of passive immunisation (via MV or mAb) to prevent RSV disease after birth in 72 Gavi-eligible countries was previously estimated by Li et al. [15] using a static model called McMarcel (Multi-Country Model Application for RSV Cost-Effectiveness poLicy), made available as an R package [33]. This model used country-specific estimates of total RSV incidence, hospitalisation probabilities and hospital and community case fatality ratios (CFR) based on the Shi et al. meta-analysis [3] of global RSV burden. These estimates were used in a probabilistic sensitivity analysis where incidence and CFR values are generated from distributions around the age-specific mean values. The disease burden and the associated costs were then reduced proportionally to the assumed efficacy of MV and mAb, while also calculating the cost of the intervention. Since all measures are probabilistically treated, the model produces a distribution for the cost effectiveness of the interventions as well.

In this study, we adopted the McMarcel analytical framework, but modified it to incorporate observed age-specific incidence rates, and treatment costs of RSV disease from Kenya and South Africa, while also distinguishing disease episodes by whether they involved medical treatment.

For South Africa, we estimated the cost of outpatient care (25 USD, CI95: 18.3-31.8) and the cost of hospitalisation by age group (SI Figure 1) **(Moyes et al. 2022)**. Hospitalisation costs include facility fees and the cost of consultations, comprising approximately 60-80% of the total cost, while in age groups where ICU visits were observed, they comprise approximately 20% of the total in-patient cost. Approximately 4 children under 5 years of age were enrolled during RSV seasons (2 outside the season) per week at each site for cost estimation. Frequency of ICU visits and lengths of stay were averaged in 3-month age bands. In South Africa hospital care for infants is free of charge, therefore costs of RSV disease requiring hospitalisation are largely absorbed by the healthcare system. In the case of Kenya, only the total cost to the healthcare system at the Siaya site (Siaya County Referral Hospital) was available. We used costs from Siaya because at the other available site (Kilifi), costs are largely subsidised from international sources and therefore do not reflect typical costs to the healthcare system. We used the ratio of inpatient to outpatient costs (per episode) of RSV-associated illness from Malawi [34], which was 4.9, and the number of inpatient and outpatient episodes to estimate inpatient and outpatient costs separately from the total cost. In Kenya, a large proportion of the total cost of hospitalisation falls on households. These costs comprise both direct out-of-pocket payments for drugs and transportation and loss of income due to absence from work. In the cost-effectiveness analysis we summed the costs to the healthcare system and those to households (SI Table 2) to calculate the cost of disease burden and its reduction by interventions.

#### Fitting efficacy estimates and using different models of efficacy

For the base case, we updated previous efficacy estimates [15] using recent results of clinical trials of the maternal vaccine ResVax [6] and nirsevimab [12], fitting to the 95% credible intervals and mean values of efficacy against symptomatic disease, hospitalisation and severe disease using beta distributions to reflect uncertainty around mean efficacy (SI Methods). The skewed confidence intervals of the MV efficacy against severe disease resulted in a fit with a lower mean than reported in the clinical trial (SI Table 4, SI Methods), and can therefore be considered a conservative estimate.

There are several new MV candidates in development and interim results recently became available suggesting efficacy against both all symptomatic and severe disease to be over 80% [13], although reported confidence intervals were very large. We used these interim efficacy values in a scenario sensitivity analysis for MV.

In the base case, we assumed that both MV and mAb have a fixed duration of protection (3 months for MV and 5 months for mAb) and to immediately fall to zero beyond that point. We performed a scenario sensitivity analysis with regard to this assumption by assuming that efficacy decays exponentially over time, with the same average value as observed in the clinical trials over the 0-90 day (MV) and 0-150 day (mAb) periods and half-life values identical or as close as possible to the reported values (SI Table 2 and 3, SI Methods). Exponential waning, therefore, implies that efficacy in the earliest life period is the highest and gradually wanes with time.

#### Probabilistic sensitivity analysis

Gamma distributions were fitted to 95% CIs of RSV incidence from both countries to reflect the uncertainty of estimates. Our dataset distinguishes between non-severe (ARI for Kenya and ILI for South Africa, SI Table 1) and severe cases on the one hand and medically attended and not attended cases on the other. In addition, we also estimate community incidence of severe cases, inferred from the hospital-based incidence via estimates of healthcare-seeking behaviour.

In a probabilistic sensitivity analysis, 5000 samples were generated per age group from the distributions of the uncertain input parameters. These rates were then multiplied by health utility measured in disability-adjusted life years (DALYs) for non-severe and severe disease (SI Table 2) to calculate the total disease burden. Similarly, incidence rates were multiplied by unit cost estimates to calculate total medical costs.

We first calculated mean and median values with corresponding credible intervals (50%, 95%) for disease burden in terms of case numbers (non-hospitalised and hospitalised cases, deaths) and in units of DALYs, as well as the costs before the interventions.

In the next step, we calculated the effect of interventions by using the distributions generated from efficacy figures and computing the number of cases (non-hospitalised, hospitalised and deaths) averted, DALYs averted and the medical cost averted. As the final measure of cost-effectiveness, we computed the incremental cost of the intervention (intervention cost minus medical costs averted) and the incremental cost per DALY averted (ICER) of each intervention compared to no intervention. We used a 3% discounting rate both for health outcomes and costs, and costs are in units of 2020 US dollars (USD). WHO-UNICEF estimates of national immunization coverage (WEUNIC) for the Bacillus Calmette– Guérin (BCG) vaccine in 2016 was used as a proxy for both interventions (MV and mAb) [15] (86% in Kenya and 95% in South Africa).

As a metric of cost-effectiveness we took the ratio of the incremental cost to DALYs averted (ICER, incremental cost-effectiveness ratio). To estimate the cost-effectiveness of interventions we used three different assumptions for the price of a complete course of MVs (3, 10, 30 USD) and mAbs (6, 20, 60 USD). The lowest prices used were the baseline assumptions in the Li et al. [15] cost-effectiveness study on RSV interventions in LMICs, whereas prices above 30 and 60 USD for MV and mAB, respectively, lead to median ICERs exceeding the GDP per capita of the two countries and are therefore unaffordable.

## Ethics statement for data use

The study received an ethics approval under the name ‘*Cost effectiveness analysis for RSV disease interventions in Kenya and South Africa*’ from the Observational Research Ethics Committee of the *London School of Hygiene & Tropical Medicine*, LSHTM Ethics Reference number: 22670. Local ethical approvals for this study’s analyses were obtained from KEMRI (Kenya; reference number SERU 3939) and the University of the Witwatersrand, Johannesburg (South Africa; reference number M121195), respectively. This activity was reviewed by CDC and was conducted consistent with applicable federal law and CDC policy [Project ID: 0900f3eb81f6f11c]

## Results

### Disease burden impact

The total DALY burden is dominated by deaths. In Kenya, approximately 97% of the total DALYs (Figure 4) are due to life years lost because of death, while the figure is approximately 95% for South Africa.

Our incidence data shows a concentration of deaths in the first 6 months of life. In Kenya 27% of RSV-associated deaths are estimated to be in the first 3 months of life and 44% in the first 6 months (Figure 3). In South Africa 44.5% of RSV-associated deaths are estimated to be in the first 3 months of life and 71% in the first 6 months.

There is a similar concentration for severe disease (SARIs). In both countries we observed peak incidence at 1 month of age for SARI episodes (Figure 1 and 2). Out of all SARI episodes (under 5 year of age) in South Africa, 48% are infants in their first 3 months of life and 62% in their first 6 months. For Kenya, the corresponding figures are 45% and 65%.

**Figure 2.**
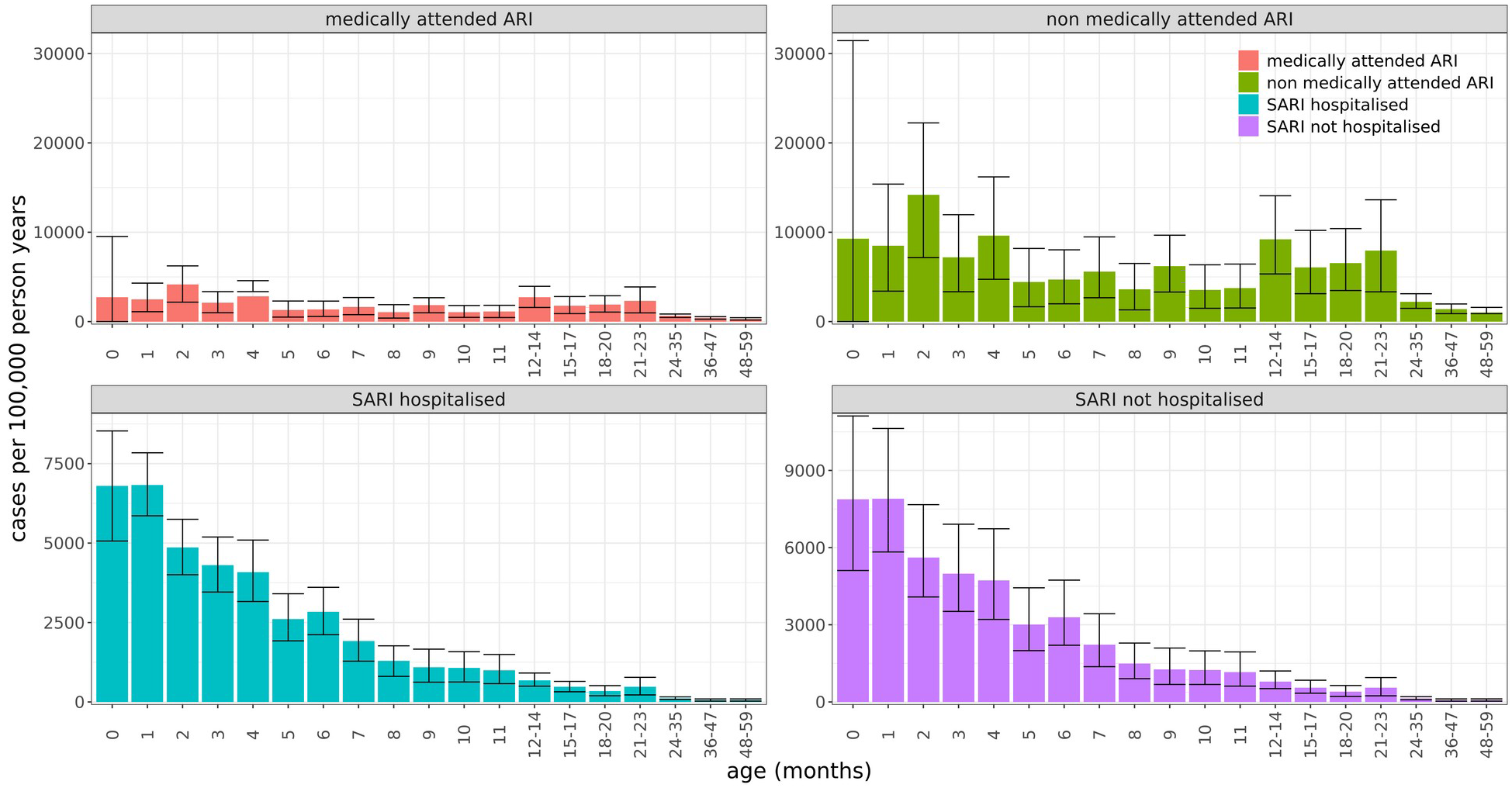
RSV-associated ARI and SARI rates per 100,000 person-years in South Africa by age group and medical status. ARI rates shown are before applying the adjustment described in Methods that aligns the South African ILI rates with the ARI definition.

As a consequence of this relative concentration of severe cases and deaths in the first months of life where both MV or mAb have their effect, the interventions can lead to a substantial reduction in disease burden.

Assuming a 3-month duration of effectiveness, we estimate that MV in Kenya would reduce the incidence of RSV hospitalisations and deaths by 14% (CI95: 11-17%) and 9% (CI95: 2-16%), respectively, using the efficacy figures from the Novavax Phase III trial [7].

Giving mAb to all newborns in Kenya, and assuming a 5-month duration of the effect, will reduce the incidence of RSV hospitalisation among children aged <5 years by 33% (CI95: 28-37.5%) and RSV-associated deaths by 28% (CI95: 23-33%) (Figure 3).

**Figure 3.**
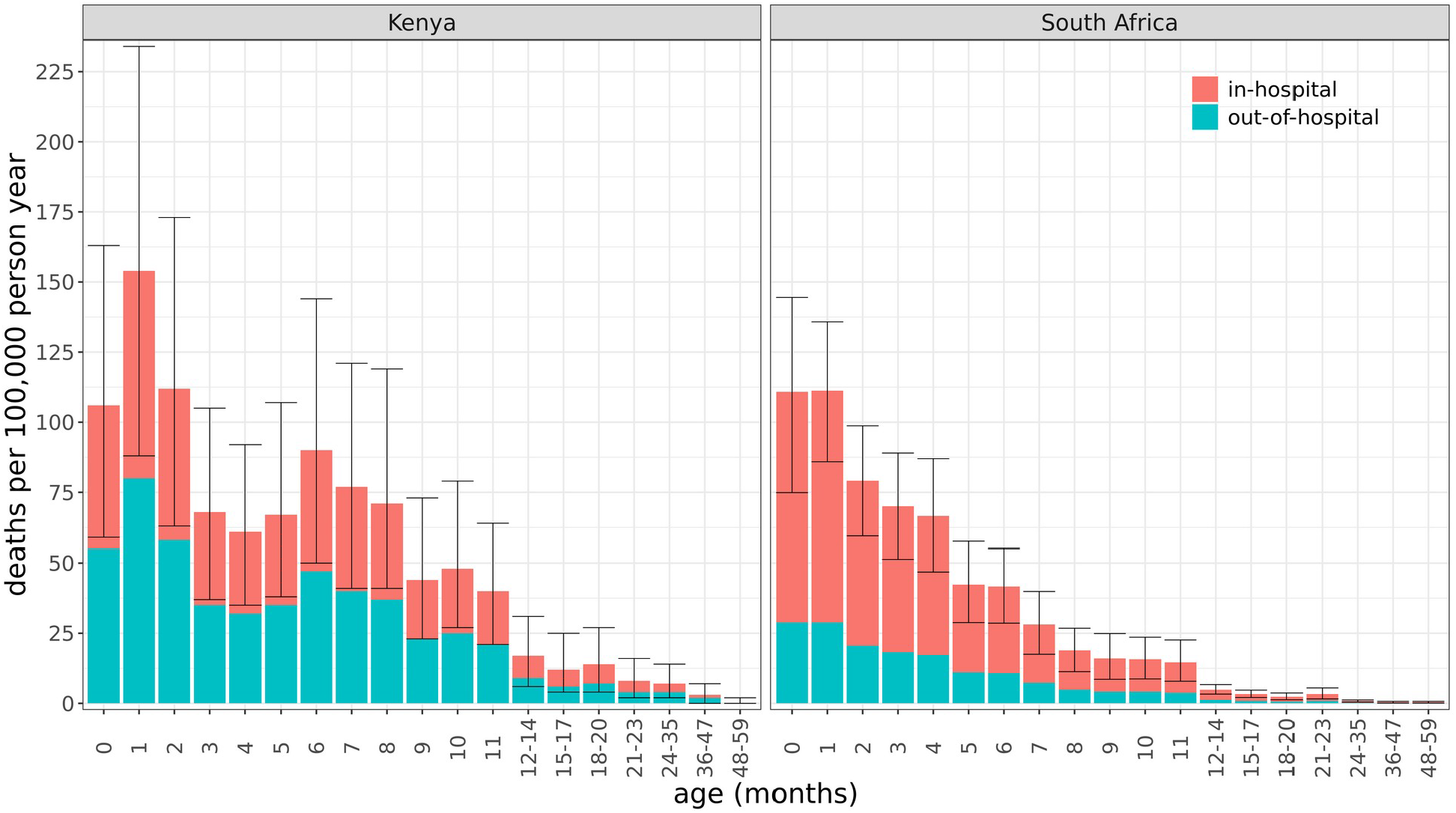
RSV-associated deaths in Kenya and South Africa per 100,000 person-years

For South Africa, hospitalisations are reduced by 17% (13-21%) and deaths by 14% (CI95: 4-25%) for MV. For mAb the reduction is 43% (CI95: 36-48%) for hospitalisations and 48% (CI95: 41-54%) for deaths. The reduction in deaths is higher than in Kenya due to a stronger concentration of RSV-associated deaths in the first months of life.

Correspondingly, we estimate a reduction of total disease burden (DALYs) of 9% (MV, CI95: 2-15%) and 25% (mAb, CI95: 21-30%) for Kenya, whereas, for South Africa, the corresponding figures are 14% (MV, CI95: 4-24%) and 47% (mAb, CI95: 40-53%).

### Cost-effectiveness

With the exception of a MV campaign with a dose price of 30 USD (which raises the median ICER to 4524 for Kenya and 4694 USD for South Africa) the median incremental cost per (undiscounted) DALY averted (incremental cost-effectiveness ratio or ICER) of the interventions is below 3000 USD per DALY averted for all scenarios (the highest value is 2650 USD). (Figure 5).

Using discounted DALYs leads to median ICERs of approximately 6000 USD for mAb for the highest dose price of 60USD, while the median ICERs for MV at 30 USD rise to approximately 10,000 USD for both countries. These values result from the relatively low MV efficacy estimates from the Novavax trial [7].

In South Africa, because of the large reduction in hospitalised cases and thereby total averted medical costs (Figure 4c), both interventions become cost-saving for the lowest dose price (Figure 5). The incremental cost for MV falls to −4 million USD (CI95: −6.4 to −1.9 million) at the lowest dose price and in the case of mAb to −10.7 million USD (−14.3 to −7.3 million). At the highest dose prices of MV in contrast the median ICER is 2641 USD (CI95: 2153-3291) for MV (30 USD per dose) and 4694 USD (CI95: 2650 - 16362) at 60 USD for mAb in South Africa.

**Figure 4.**
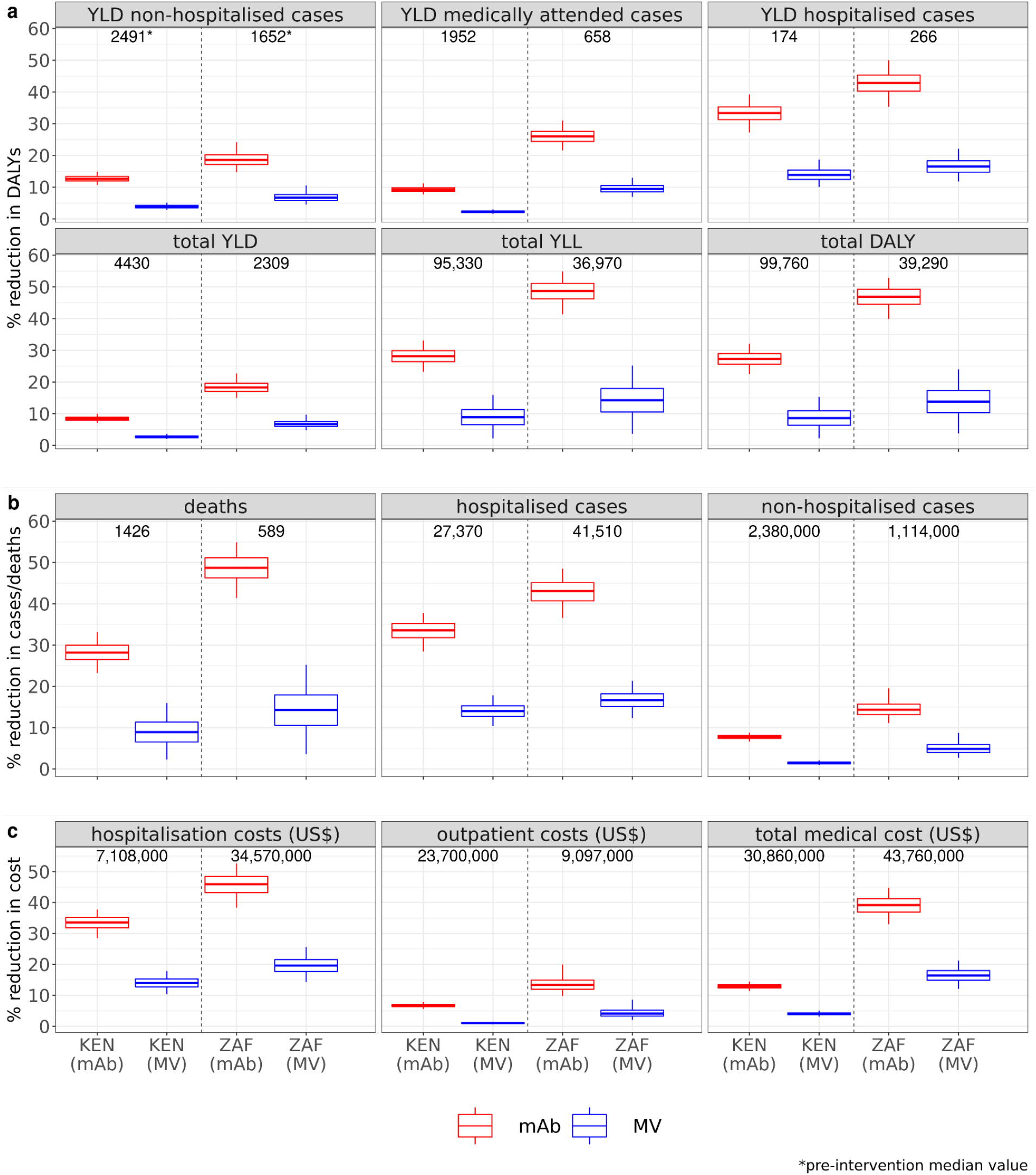
**a**. Percentage reduction (y axis) in non-hospitalised and hospitalised RSV cases and RSV-associated deaths in response to maternal vaccination or monoclonal antibody use. Solid horizontal lines show the reduction as a percentage of the variables’ pre-intervention median values, which are displayed by the numbers at the top of panels (the pre-intervention values are the same for MV and mAb). Horizontal lines show the median values. Boxes show 50% and whiskers the 95% credible intervals. **b**. Percentage reduction in subtypes of disease burden (DALYs) in response to maternal vaccination or monoclonal antibody campaigns. **c**. Percentage reduction in costs of disease burden (USD) in response to maternal vaccination or monoclonal antibody campaigns.

**Figure 5:**
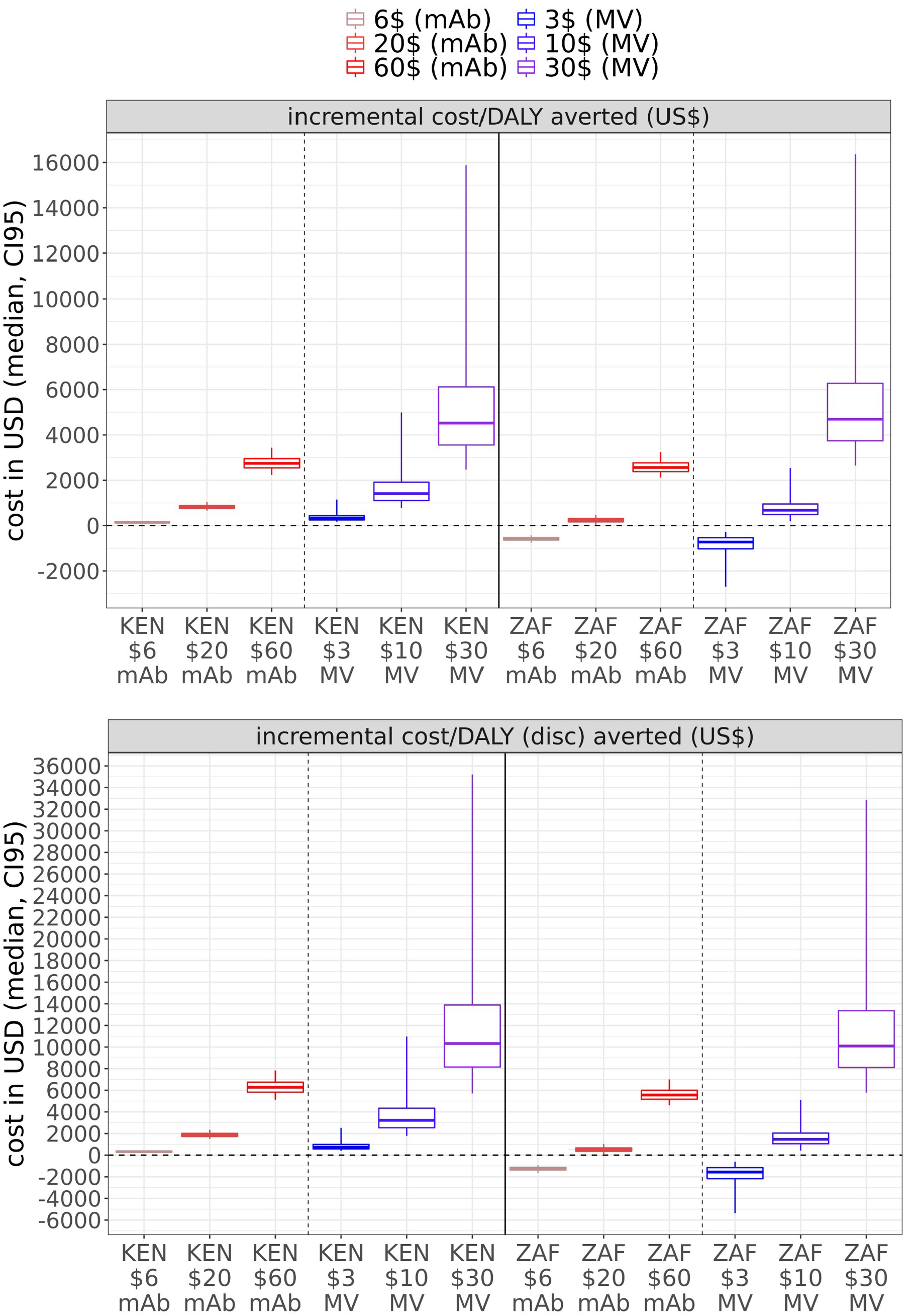
Cost-effectiveness of maternal vaccination and monoclonal antibody campaigns at three different levels of dose prices in Kenya (KEN) and South Africa (ZAF), expressed as incremental cost/DALY averted, using **a)** non-discounted and **b)** discounted DALYs. Negative values represent net savings. Horizontal lines show the median values. Boxes show 50%, whiskers the 95% credible intervals.

For Kenya, the median (undiscounted) ICERs at the lowest dose prices are 142 USD (108-193) for mAB and 321 USD (175-1148) for MV, or, using discounted DALYs, 325 USD for mAb (CI95: 246-440) and 734 for MV (CI95: 402-2518). At the highest dose prices the median (undiscounted) ICER rises slightly above 2500 USD for mAB (2748 USD) and to 4525 USD for MV (CI95: 2470 - 15881).

### Sensitivity analysis

The calculated ICER values above are sensitive to efficacy estimates since the proportion of the disease burden averted depends on the efficacy of the antibodies or vaccine used. Using the interim efficacy data [13] for the Pfizer MV (SI Table 4) reduces median (undiscounted) ICER values below 2000 USD for all MV scenarios (SI Figure 2).

Similarly, an exponential waning model implies some level of protection lasting beyond the defined duration of protection and also leads to median ICERs lower than in the default scenarios (SI Figure 3), with all median ICERs below 3500 USD, and with the exception of a MV dose price of 30 USD also below 3000 USD. This result strongly depends on assumptions about the half-life values (SI Table 3 and SI Methods).

Using the narrower ILI definition for non-severe disease in South Africa does not substantially alter the estimate for total disease burden. While years lived with disability (YLD) would be lower in this case, the total burden in DALYs only changes by approximately 1% since it is dominated by deaths.

## Discussion

Because most RSV-associated SARI cases and a large proportion of deaths occur in the first 3 months of life according to our hospital surveillance data in both Kenya and South Africa, mAb or MV programmes against RSV disease may be more cost-effective than suggested by previous analyses based on estimates of community incidence [1]–[3]. Whereas in previous studies, it was assumed that severe disease and death have a similar age profile as non-severe RSV infections, our incidence data suggests severe disease is more concentrated in the first months of life, potentially increasing the impact of these interventions on DALYs, and hence making them more cost-effective.

Deaths dominate the total DALY burden of RSV disease for infants. Phase IIb data from recent trials for a new mAb candidate (nirsevimab) show 78% efficacy against severe cases. However, even ResVax, a MV with reported lower efficacy (mean 48.3%) against severe disease has a strong effect on DALYs.

Since most of the total disease burden measured in DALYs (Figure 3b) is due to deaths, the main driver of cost-effectiveness of the intervention is its impact on deaths. The expected reduction in deaths, in the range of 8% (MV) to 26% (mAb) for Kenya and 14% (MV) to 48% (mAb) for South Africa, leads to an almost equivalent reduction in total disease burden. Additionally, since medical costs are dominated by hospitalisations associated with SARIs, the comparable reduction in hospitalisations leads to a similar reduction in RSV-associated medical costs. As a combined effect of these two, we arrive at median estimates of (undiscounted) ICERs below 2800 USD for all dose prices except for the highest price (30 USD) in the case of MV. For dose prices below 30USD for MV and below 60 USD for mAb median ICERs are below 2000 USD, and in the case of South Africa interventions are cost-saving for the lowest dose price (3 USD for MV and 6 USD for mAb).

Our analysis uses published phase III results from a RSV MV (ResVax) and phase II results from a next-generation RSV mAb (nirsevimab). We find that both biologics lead to interventions with a median ICER below 2800 USD at a cost per course of 3 to 10 USD for MV and 6 to 60 USD for mAb, respectively. At a MV dose price of 30 USD the median ICER rises to approximately 4500 USD in Kenya and to nearly 5000 USD in South Africa. We note this result will be sensitive to the distribution used to fit the currently available [7] efficacy data of the MV (ResVax), with interim results suggesting higher efficacy for other vaccine candidates [13]. However, neither product is currently available on the market. In particular, ResVax did not meet its primary clinical endpoint in the United States pre-specified by the Food and Drug Administration (lower bound of the 97.52% confidence interval ≥30% efficacy against medically significant LRTI up to 90 days of life), although it showed significant efficacy against RSV-associated LRTI hospitalisation and RSV LRTI with severe hypoxemia (on average 44.4% and 48.3%, respectively). Also, more than half of the participants enrolled in the clinical trial were in South Africa and efficacy figures in low- and middle-income countries were significantly higher, especially when comparing efficacies against hospitalisation and at the later (180 days after birth) point estimates [6]. While we used the more conservative trial-wide efficacy estimates in this study, these results suggest potentially higher efficacy in a low and middle-income setting and, therefore even improved cost-effectiveness. While ResVax does not have a clear path to licensure and use, our analyses using the published estimates suggest that it could be lifesaving and its incremental cost per DALY averted below 2000 USD at lower dose prices (10USD for MV and 20 USD for mAb) in Kenya and South Africa.

Our study has several limitations. We did not consider a scenario where interventions are given before and during RSV seasons rather than throughout the whole year. Seasonal immunisation campaigns are likely to be more efficient (per dose) [35] in countries with a clear seasonal RSV pattern. On a national level, Kenya does not have a clear RSV season pattern [36], although some of its regions do, but even these regions have more spread-out RSV circulation than temperate countries [37], therefore it is not clear if seasonal campaigns would be viable as opposed to nationwide year-round immunisation. A seasonal approach could be more viable in South Africa as it has a more seasonal pattern nationally [38], [39] although with substantial variation in the timing of the season’s end. The concentration of SARI illness episodes and deaths we observed in children under 6 months of age in Kenya and South Africa, which is crucial for the favourable cost-effectiveness estimates we obtained, appears to be different from the age distribution observed in some other African countries [40]. Some incidence rates for regions in Kenya (which were then combined to a national estimate) had to be projected from the areas of data collection using ratios of health-seeking behaviour. More sites for data collection would further support the reliability of these estimates. Further studies would be important to investigate if this concentration of severe RSV disease in early infancy is a more general phenomenon in African countries or perhaps specific to the two countries analysed in the current study. Finally, there is uncertainty about the absolute levels and duration of efficacy of both MV and mAb as several candidates are still in clinical trials. The MV candidate (ResVax) [6] that completed phase III trials had different results in the United States and LMICs. As results from clinical trials of other MV candidates are published the cost-effectiveness estimates might improve further, as suggested by higher efficacy figures from interim results of a MV candidate [13].

In summary, our results suggest that public health interventions based on vaccination or monoclonal antibodies against RSV disease in infants in two African countries might be cost-effective, depending on the country’s willingness-to-pay values. Specifically, for maternal vaccine dose prices up to 10 USD we estimated the (median) incremental cost per (undiscounted) DALY averted to be below 1500 USD, and for dose prices of monoclonal antibodies up to 20 USD to be below 900 USD. As recent estimates of cost-effectiveness thresholds based on health opportunity costs [41] tend to fall below previously commonly used thresholds which were in the range of 1-3x of GDP per capita, the new MV candidates’ efficacy (as well as their effectiveness following implementation) might be crucial to making decisions, especially in the case of MV in Kenya. In the coming years, further analysis and planning for these interventions in the light of multiple vaccine and antibody candidates currently in Phase III clinical trials will continue to be an important priority for public health policy in the coming years.

## Data Availability

R scripts used for the cost-effectiveness analysis and the corresponding data input files are available at https://github.com/mbkoltai/RSV-CEA-Kenya-South-Africa

## Abbreviations used

HCW: healthcare worker
LRTI: lower respiratory tract infection
HUS: healthcare utilisation survey
HMS: Household Morbidity Surveillance
DHS: Demographic and Health Surveys
MV: maternal vaccination
mAb: monoclonal antibodies

## Code accessibility

Scripts used for the cost-effectiveness analysis and the corresponding data input files are available at https://github.com/mbkoltai/RSV-CEA-Kenya-South-Africa

## Acknowledgements

We thank Elisabeth Vodicka and Clint Pecenka from PATH for coordinating the project and for their useful comments and suggestions on the analysis.

## Literature

[1] P. Obando-Pacheco et al., ‘Respiratory Syncytial Virus Seasonality: A Global Overview’, J. Infect. Dis., vol. 217, no. 9, pp. 1356–1364, Apr. 2018, doi: 10.1093/infdis/jiy056.

[2] H. Nair et al., ‘Global burden of acute lower respiratory infections due to the respiratory syncytial virus in young children: a systematic review meta-analysis’, The Lancet, vol. 375, no. 9725, pp. 1545–1555, May 2010, doi: 10.1016/S0140-6736(10)60206-1.

[3] T. Shi et al., ‘Global, regional, and national disease burden estimates of acute lower respiratory infections due to respiratory syncytial virus in young children in 2015: a systematic review and modelling study’, The Lancet, vol. 390, no. 10098, pp. 946–958, Sep. 2017, doi: 10.1016/S0140-6736(17)30938-8.

[4] K. L. O’Brien et al., ‘Causes of severe pneumonia requiring hospital admission in children without HIV infection from Africa and Asia: the PERCH multi-country case-control study’, The Lancet, vol. 394, no. 10200, pp. 757–779, Aug. 2019, doi: 10.1016/S0140-6736(19)30721-4.

[5] ‘RSV Vaccine and mAb Snapshot’. https://www.path.org/resources/rsv-vaccine-and-mab-snapshot/ (accessed Jul. 06, 2021).

[6] S. A. Madhi et al., ‘Respiratory Syncytial Virus Vaccination during Pregnancy and Effects in Infants’, N. Engl. J. Med., vol. 383, no. 5, pp. 426–439, Jul. 2020, doi: 10.1056/NEJMoa1908380.

[7] ‘Respiratory Syncytial Virus Vaccination during Pregnancy and Effects in Infants | NEJM’. https://www.nejm.org/doi/full/10.1056/NEJMoa1908380 (accessed Nov. 15, 2021).

[8] ‘GSK starts phase 3 study of RSV maternal candidate vaccine | GSK’. https://www.gsk.com/en-gb/media/press-releases/gsk-starts-phase-3-study-of-rsv-maternal-candidate-vaccine/ (accessed Jun. 10, 2021).

[9] GlaxoSmithKline, ‘ A Phase III, Randomized, Multi-country Study to Evaluate the Lot-to-lot Consistency of GSK’s Investigational RSV Maternal Vaccine and the Immune Response, Safety and Reactogenicity of RSV Maternal Vaccine When Co-administered With GSK’s Quadrivalent Influenza D-QIV Vaccine in Healthy Non-pregnant Women 18-49 Years of Age.’, clinicaltrials.gov, Clinical trial registration NCT05045144, Dec. 2021. Accessed: Feb. 17, 2022. [Online]. Available: https://clinicaltrials.gov/ct2/show/NCT05045144

[10] Pfizer, ‘A PHASE 3, RANDOMIZED, DOUBLE-BLINDED, PLACEBO-CONTROLLED TRIAL TO EVALUATE THE EFFICACY AND SAFETY OF A RESPIRATORY SYNCYTIAL VIRUS (RSV) PREFUSION F SUBUNIT VACCINE IN INFANTS BORN TO WOMEN VACCINATED DURING PREGNANCY’, clinicaltrials.gov, Clinical trial registration NCT04424316, Aug. 2021. Accessed: Aug. 10, 2021. [Online]. Available: https://clinicaltrials.gov/ct2/show/NCT04424316

[11] ‘Nirsevimab shows positive topline results in RSV Phase 2/3 MEDLEY trial’. https://www.sanofi.com/media-room/press-releases/2021/2021-04-2608-00-002216474 (accessed Jun. 10, 2021).

[12] M. P. Griffin et al., ‘Single-Dose Nirsevimab for Prevention of RSV in Preterm Infants’, N. Engl. J. Med., vol. 383, no. 5, pp. 415–425, Jul. 2020, doi: 10.1056/NEJMoa1913556.

[13] ’RSVVW’21 conference, Talk by Kimberly J. Center (Pfizer)’, RESVINET, Nov. 10, 2021. https://www.resvinet.org/rsvvw21.html (accessed Feb. 03, 2022).

[14] C. O. I. D. and B. G. Committee, ‘Updated Guidance for Palivizumab Prophylaxis Among Infants and Young Children at Increased Risk of Hospitalization for Respiratory Syncytial Virus Infection’, Pediatrics, vol. 134, no. 2, pp. e620–e638, Aug. 2014, doi: 10.1542/peds.2014-1666.

[15] X. Li, L. Willem, M. Antillon, J. Bilcke, M. Jit, and P. Beutels, ‘Health and economic burden of respiratory syncytial virus (RSV) disease and the cost-effectiveness of potential interventions against RSV among children under 5 years in 72 Gavi-eligible countries’, BMC Med., vol. 18, no. 1, p. 82, Dec. 2020, doi: 10.1186/s12916-020-01537-6.

[16] G. O. Emukule et al., ‘The Burden of Influenza and RSV among Inpatients and Outpatients in Rural Western Kenya, 2009–2012’, PLOS ONE, vol. 9, no. 8, p. e105543, Aug. 2014, doi: 10.1371/journal.pone.0105543.

[17] J. A. Fuller et al., ‘Estimation of the National Disease Burden of Influenza-Associated Severe Acute Respiratory Illness in Kenya and Guatemala: A Novel Methodology’, PLOS ONE, vol. 8, no. 2, p. e56882, Feb. 2013, doi: 10.1371/journal.pone.0056882.

[18] J. A. Dawa et al., ‘National burden of hospitalized and non-hospitalized influenza-associated severe acute respiratory illness in Kenya, 2012-2014’, Influenza Other Respir. Viruses, vol. 12, no. 1, pp. 30–37, Jan. 2018, doi: 10.1111/irv.12488.

[19] G. Bigogo, A. Audi, B. Aura, G. Aol, R. F. Breiman, and D. R. Feikin, ‘Health-seeking patterns among participants of population-based morbidity surveillance in rural western Kenya: implications for calculating disease rates’, Int. J. Infect. Dis., vol. 14, no. 11, pp. e967–e973, Nov. 2010, doi: 10.1016/j.ijid.2010.05.016.

[20] D. R. Feikin et al., ‘The Burden of Common Infectious Disease Syndromes at the Clinic and Household Level from Population-Based Surveillance in Rural and Urban Kenya’, PLOS ONE, vol. 6, no. 1, p. e16085, Jan. 2011, doi: 10.1371/journal.pone.0016085.

[21] ’The DHS Program - Kenya: Standard DHS, 2014 Dataset’. https://www.dhsprogram.com/data/dataset/Kenya_Standard-DHS_2014.cfm?flag=0 (accessed Jun. 30, 2021).

[22] A. A. Tazinya, G. E. Halle-Ekane, L. T. Mbuagbaw, M. Abanda, J. Atashili, and M. T. Obama, ‘Risk factors for acute respiratory infections in children under five years attending the Bamenda Regional Hospital in Cameroon’, BMC Pulm. Med., vol. 18, no. 1, p. 7, Jan. 2018, doi: 10.1186/s12890-018-0579-7.

[23] J. A. G. Scott et al., ‘Profile: The Kilifi Health and Demographic Surveillance System (KHDSS)’, Int. J. Epidemiol., vol. 41, no. 3, pp. 650–657, Jun. 2012, doi: 10.1093/ije/dys062.

[24] C. Cohen et al., ‘In- and Out-of-hospital Mortality Associated with Seasonal and Pandemic Influenza and Respiratory Syncytial Virus in South Africa, 2009–2013’, Clin. Infect. Dis., vol. 66, no. 1, pp. 95–103, Jan. 2018, doi: 10.1093/cid/cix740.

[25] S. Tempia et al., ‘Quantifying How Different Clinical Presentations, Levels of Severity, and Healthcare Attendance Shape the Burden of Influenza-associated Illness: A Modeling Study From South Africa’, Clin. Infect. Dis., vol. 69, no. 6, pp. 1036–1048, Aug. 2019, doi: 10.1093/cid/ciy1017.

[26] K. K.-L. Wong et al., ‘Healthcare utilization for common infectious disease syndromes in Soweto and Klerksdorp, South Africa’, Pan Afr. Med. J., vol. 30, no. 271, Art. no. 271, Aug. 2018, doi: 10.11604/pamj.2018.30.271.14477.

[27] ’Healthcare utilisation patterns for respiratory and gastrointestinal syndromes and meningitis in Msunduzi municipality, Pietermaritzburg, KwaZulu-Natal Province, South Africa, 2013 | McAnerney | South African Medical Journal’. http://www.samj.org.za/index.php/samj/article/view/12596 (accessed Jun. 30, 2021).

[28] I. Rudan, C. Boschi-Pinto, Z. Biloglav, K. Mulholland, and H. Campbell, ‘Epidemiology and etiology of childhood pneumonia’, Bull. World Health Organ., vol. 86, no. 5, pp. 408–416, May 2008, doi: 10.2471/blt.07.048769.

[29] J. Murray et al., ‘Determining the Provincial and National Burden of Influenza-Associated Severe Acute Respiratory Illness in South Africa Using a Rapid Assessment Methodology’, PLOS ONE, vol. 10, no. 7, p. e0132078, Jul. 2015, doi: 10.1371/journal.pone.0132078.

[30] Statistics South Africa and South Africa, Eds., South Africa Demographic and Health Survey 2016. Key indicators report. Pretoria: Statistics South Africa, 2017.

[31] ‘Case definitions’. https://www.who.int/teams/control-of-neglected-tropical-diseases/yaws/diagnosis-and-treatment/global-influenza-programme (accessed Aug. 11, 2021).

[32] J. U. Nyiro et al., ‘Surveillance of respiratory viruses in the outpatient setting in rural coastal Kenya: baseline epidemiological observations’, Wellcome Open Res., vol. 3, p. 89, Jul. 2018, doi: 10.12688/wellcomeopenres.14662.1.

[33] L. Willem, L. Xiao, M. Antillon, J. Bilcke, M. Jit, and P. Beutels, Multi-Country Model Application for RSV Cost-Effectiveness Policy (McMarcel). Zenodo, 2020. doi: 10.5281/zenodo.3663447.

[34] R. Baral et al., ‘Estimating the Economic Impact of Respiratory Syncytial Virus and Other Acute Respiratory Infections Among Infants Receiving Care at a Referral Hospital in Malawi’, J. Pediatr. Infect. Dis. Soc., vol. 9, no. 6, pp. 738–745, Dec. 2020, doi: 10.1093/jpids/piaa157.

[35] Y. Li, D. Hodgson, X. Wang, K. E. Atkins, D. R. Feikin, and H. Nair, ‘Respiratory syncytial virus seasonality and prevention strategy planning for passive immunisation of infants in low-income and middle-income countries: a modelling study’, Lancet Infect. Dis., May 2021, doi: 10.1016/S1473-3099(20)30703-9.

[36] E. B. Rose et al., ‘Respiratory syncytial virus seasonality in three epidemiological zones of Kenya’, Influenza Other Respir. Viruses, vol. 15, no. 2, pp. 195–201, 2021, doi: 10.1111/irv.12810.

[37] Y. Li, D. Hodgson, X. Wang, K. E. Atkins, D. R. Feikin, and H. Nair, ‘Respiratory syncytial virus seasonality and prevention strategy planning for passive immunisation of infants in low-income and middle-income countries: a modelling study’, Lancet Infect. Dis., vol. 21, no. 9, pp. 1303–1312, Sep. 2021, doi: 10.1016/S1473-3099(20)30703-9.

[38] R. Kyeyagalire et al., ‘Hospitalizations associated with influenza and respiratory syncytial virus among patients attending a network of private hospitals in South Africa, 2007–2012’, BMC Infect. Dis., vol. 14, no. 1, p. 694, Dec. 2014, doi: 10.1186/s12879-014-0694-x.

[39] L. Staadegaard et al., ‘Defining the seasonality of respiratory syncytial virus around the world: National and subnational surveillance data from 12 countries’, Influenza Other Respir. Viruses, vol. 15, no. 6, pp. 732–741, 2021, doi: 10.1111/irv.12885.

[40] R. S. Laufer et al., ‘Cost-effectiveness of infant respiratory syncytial virus preventive interventions in Mali: A modeling study to inform policy and investment decisions’, Vaccine, vol. 39, no. 35, pp. 5037–5045, Aug. 2021, doi: 10.1016/j.vaccine.2021.06.086.

[41] J. Ochalek, J. Lomas, and K. Claxton, ‘Estimating health opportunity costs in low-income and middle-income countries: a novel approach and evidence from cross-country data’, BMJ Glob. Health, vol. 3, no. 6, p. e000964, Nov. 2018, doi: 10.1136/bmjgh-2018-000964.

[42] G. M. Bigogo et al., ‘Epidemiology of Respiratory Syncytial Virus Infection in Rural and Urban Kenya’, J. Infect. Dis., vol. 208, no. suppl_3, pp. S207–S216, Dec. 2013, doi: 10.1093/infdis/jit489.

[43] ’Thembisa Project’. https://www.thembisa.org/about (accessed Jun. 30, 2021).

[44] S. Tempia et al., ‘Attributable Fraction of Influenza Virus Detection to Mild and Severe Respiratory Illnesses in HIV-Infected and HIV-Uninfected Patients, South Africa, 2012–2016 - Volume 23, Number 7—July 2017 - Emerging Infectious Diseases journal - CDC’, doi: 10.3201/eid2307.161959.

[45] C. Cohen et al., ‘Severe Influenza-associated Respiratory Infection in High HIV Prevalence Setting, South Africa, 2009–2011 - Volume 19, Number 11—November 2013 - Emerging Infectious Diseases journal - CDC’, doi: 10.3201/eid1911.130546.

[46] D. J. Nokes et al., ‘Respiratory Syncytial Virus Infection and Disease in Infants and Young Children Observed from Birth in Kilifi District, Kenya’, Clin. Infect. Dis., vol. 46, no. 1, pp. 50–57, Jan. 2008, doi: 10.1086/524019.

[47] Y. Li et al., ‘Global, regional, and national disease burden estimates of acute lower respiratory infections due to respiratory syncytial virus in children younger than 5 years in 2019: a systematic analysis’, The Lancet, vol. 399, no. 10340, pp. 2047–2064, May 2022, doi: 10.1016/S0140-6736(22)00478-0.

[48] Kenya National Bureau of Statistics (KNBS). Kenya Demographic and Health Survey 2014. Gov Kenya 2015 https://dhsprogram.com/pubs/pdf/fr308/fr308.pdf

[49] ‘GSK provides further update on phase III RSV maternal vaccine candidate programme’ https://www.gsk.com/en-gb/media/press-releases/gsk-provides-further-update-on-phase-iii-rsv-maternal-vaccine-candidate-programme/ (accessed July 14, 2022)

[50] L. L. Hammitt et al., ‘Nirsevimab for Prevention of RSV in Healthy Late-Preterm and Term Infants’, N. Engl. J. Med., vol. 386, no. 9, pp. 837–846, Mar. 2022, doi: 10.1056/NEJMoa2110275.

