## Supplementary Information for "Estimating the cost-effectiveness of maternal vaccination and monoclonal antibodies for respiratory syncytial virus in Kenya and South Africa"

#### Supplementary Tables and Figures

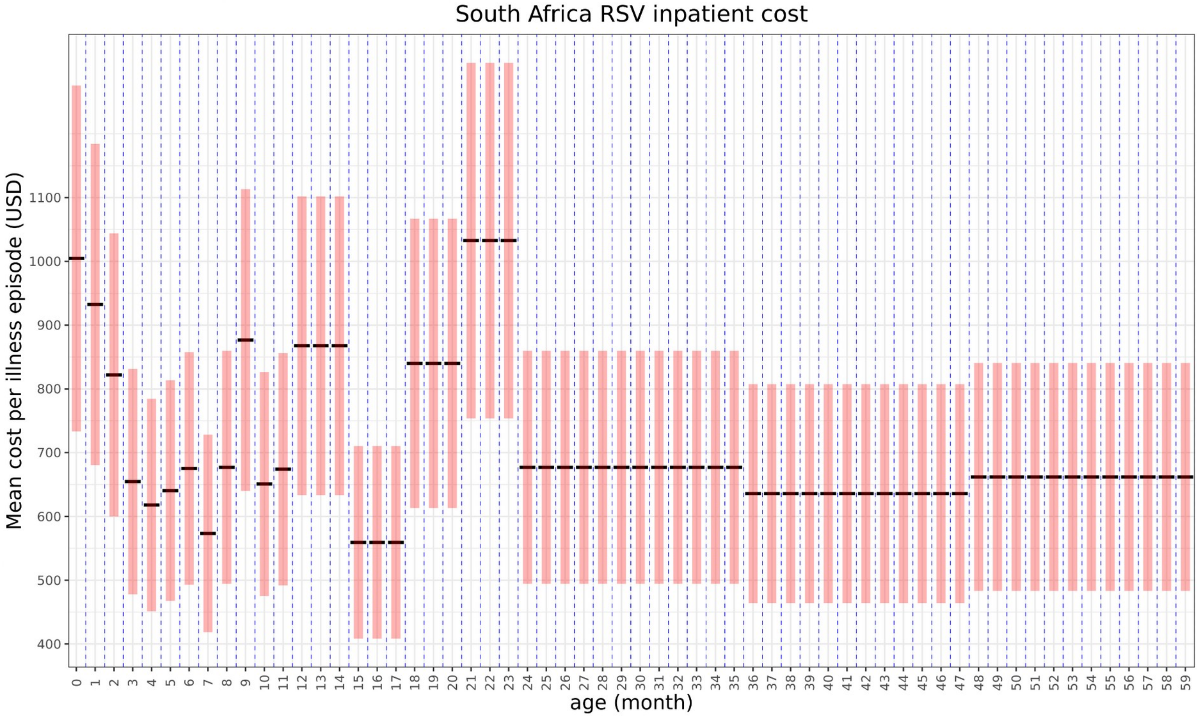


**SI Figure 1**: Cost of RSV-associated hospitalisation in South Africa by age group (mean and 95% confidence intervals). The variation above 5 months of age is mainly due to whether there are ICU cases in a given age band. Since most hospitalisations are in the first months of life this variation in older age groups does not meaningfully impact the results.

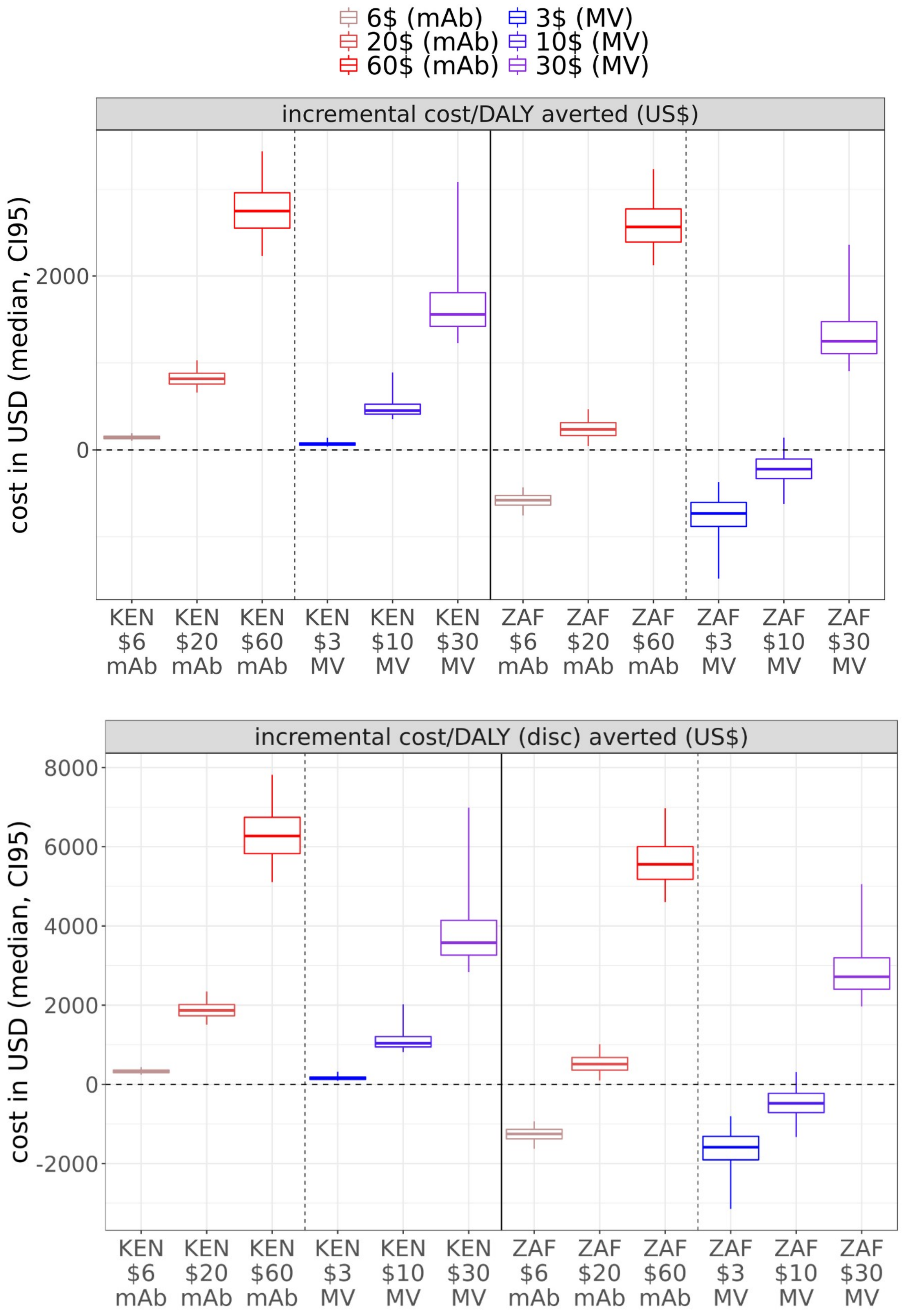


**SI Figure 2**: Incremental cost per DALY averted when using interim (higher) efficacy values for the Pfizer maternal vaccine (SI Table 4). Upper panel: undiscounted DALYs, lower panel: discounted DALYs. Horizontal lines show median values, the boxes 50% and the whiskers 95% credible intervals.

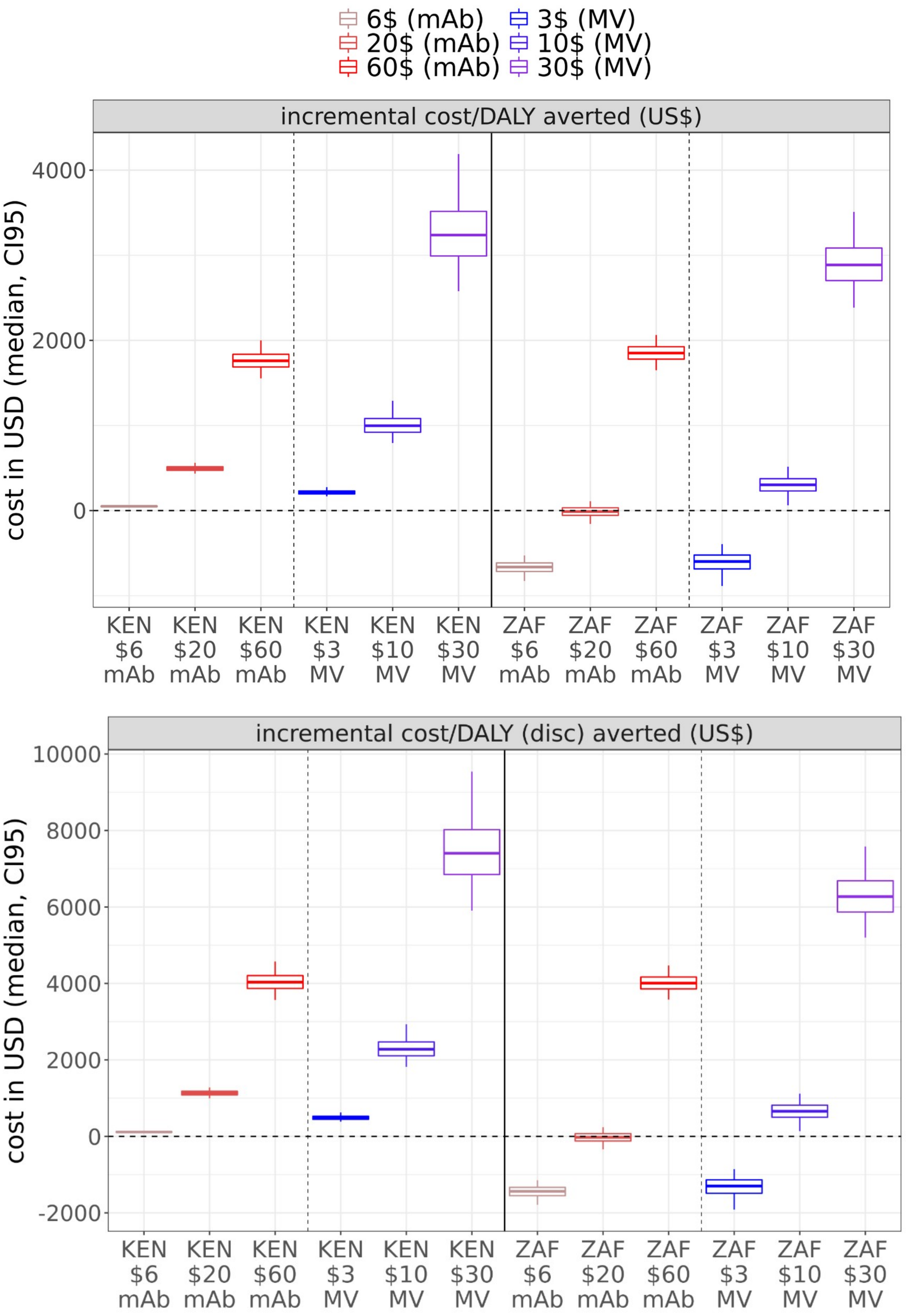


**SI Figure 3**. Incremental cost per DALY averted when assuming exponentially waning efficacy instead of a step function. Efficacy figures and parameters for exponential waning are in SI Table 2 and 3. Upper plot shows undiscounted, lower plot discounted DALYs.

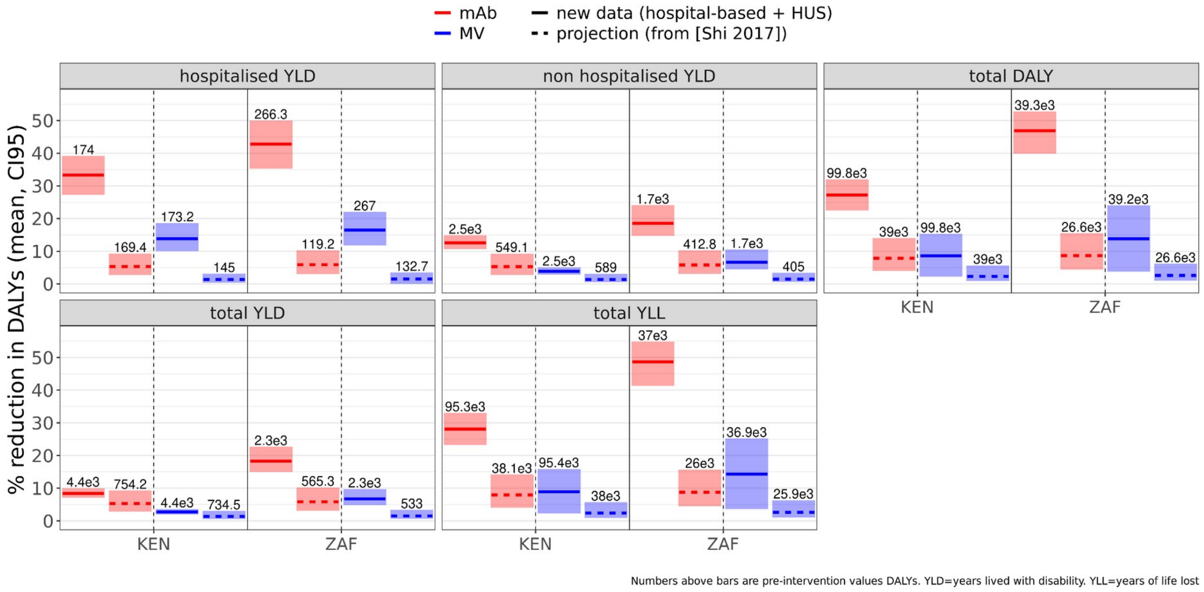


**SI Figure 4**. Reduction of disease burden in the dataset described in this paper (solid lines) compared to projections from the Shi et al. global meta-analysis [15] (dashed lines). Percentage reduction of disease burden (DALYs) in response to maternal vaccination (MV) or monoclonal antibodies (mAbs). Horizontal lines show the reduction as a percentage of the variables' pre-intervention median values, numbers above the horizontal lines show the pre-intervention median. Shaded regions show CI50s.

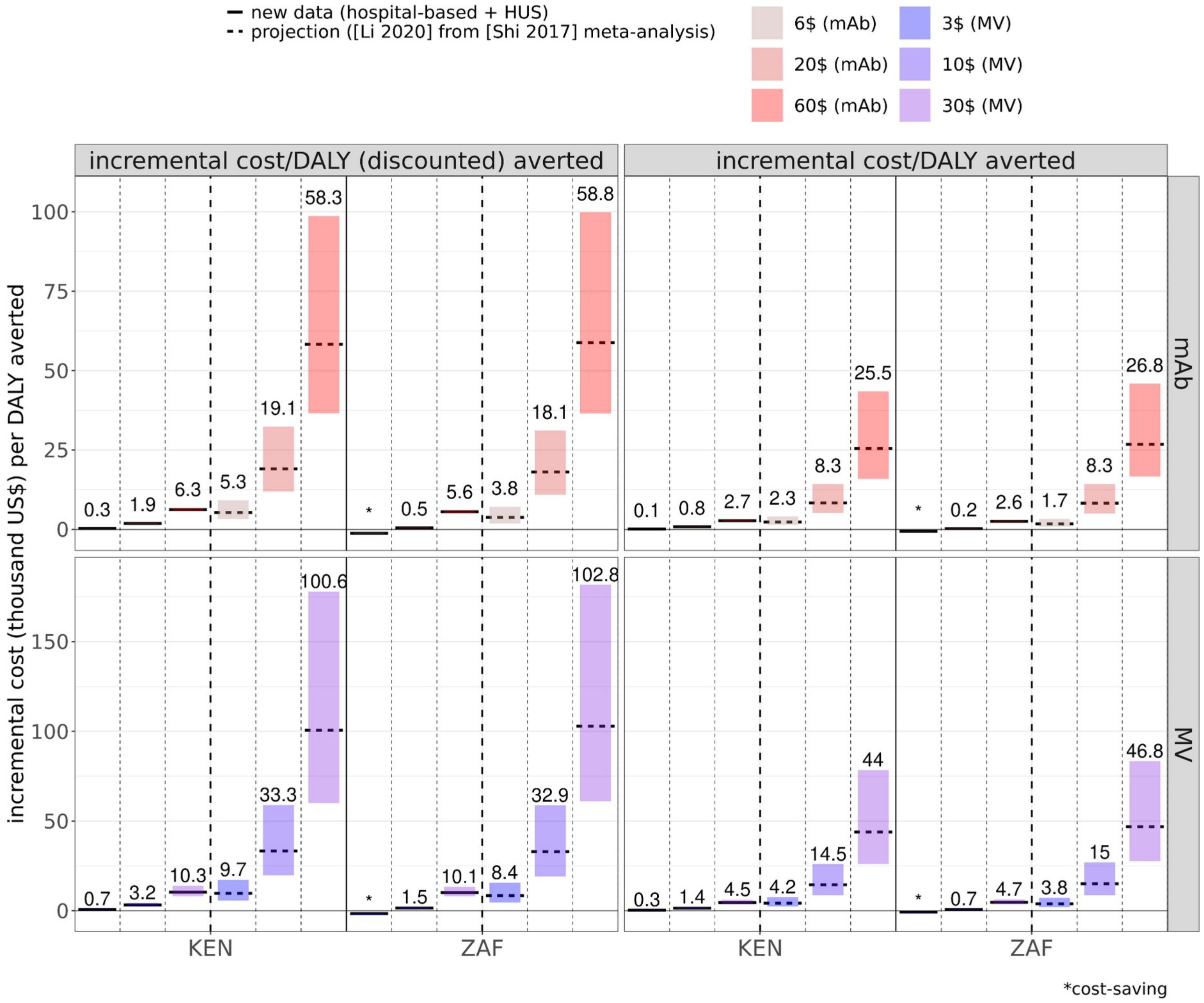

**SI Figure 5**. Incremental cost per DALY averted (median values and CI50) using the dataset described in this paper (solid lines) and calculated from previous projections from Li et al [15] (dashed lines). Efficacy values in both cases are the ones shown in SI Table 2.

**SI Table 1**: Data sources and definitions for metrics used

| Metric | Disease type | Source | Type of data | Calculation |
|---|---|---|---|---|
| $I_A$ = Rate of medically attended ARI in Asembo (base area) | ARI: acute respiratory infection.<br>Definition: acute respiratory infection (onset within 10 days) with cough, difficulty breathing, sore throat or runny nose | Kenya Medical Research Institute (KEMRI) | Hospital data from St. Elizabeth Lwak Mission Hospital (LMH) and surveillance data from Health and Demographic Surveillance System (HDSS) in Asembo, Kenya | $I_A = \frac{\frac{ARI_A}{Pop_A} * 1}{W}$<br>**ARI$_A$** = Total number of ARI cases in Asembo who visited LMH per year<br>**Pop$_A$** = Population of surveillance catchment area. The catchment area is defined in [42]. There was ongoing surveillance, so population and healthcare-seeking behavior data are up-to-date.<br>**W** = Proportion of ARI cases visiting LMH *among those who sought care*, from Household Morbidity Surveillance (HMS) [42] |
| **$I_B$** = Rate of medically attended ARI in the base region | ARI | KEMRI | Projected from rate of medically attended ARIs in Asembo by :<br>- health seeking behavior of ARI cases in Asembo from Household Morbidity Surveillance (from HMS [42])<br>- Proportion of ARI cases seeking care in Nyanza region (from DHS [21]) | $I_B = I_A \times \frac{DHS_B}{HMS_A}$<br>**DHS$_B$** = Proportion of ARI cases seeking care in base region from DHS [21]<br>**HMS$_A$** = Proportion of ARI cases seeking care, in Asembo (from HMS [42])<br>**DHS**: Demographic and Health Survey 2015 [21] |
| **$I_{M,Y}$** = Incidence of medically attended and non-attended | ARI | KEMRI | ARI rate for base region (Nyanza) projected by adjustment with risk factors [21][22] | $I_{M,Y} = I_B \times Adj_Y$<br>Adj$_Y$ = adjustment factor.<br>$Adj_Y = \left(1 + \sum_i \left(P_{i,Y} - P_{i,B}\right) \times \left(RR_i - 1\right)\right) \times \frac{DHS_Y}{DHS_B}$ |

| | | | | |
|---|---|---|---|---|
| ARI in region Y<br><br>**$I_{NM,Y}$** = Incidence of non-medically attended ARI in region Y | | | | **$P_{i,Y}$** =prevalence of risk factor i in region Y<br>**$P_{i,B}$** =prevalence of risk factor *i* in the base region (Nyanza province).<br>Risk prevalence estimated from [21] [22]. 1000 iterations were run for each risk factor (binomial distribution) to estimate credible intervals.<br>**$RR_i$**=Relative risk of ARI due to risk factor *i*, from [22]<br>**$DHS_Y$** = Proportion of ARI cases seeking care in region Y (from DHS)<br>$I_{NM,Y}=\left(I_{M,Y}\times\frac{1}{DHS_Y}\right)-I_{M,Y}$ |
| **$IF_{M,Y}$**=Incidence of medically attended RSV-associated ARI in region Y<br><br>**$IF_{NM,Y}$** = Incidence of non-medically attended RSV-associated ARI in region Y | ARI | KEMRI | Calculated from projected ARI rate for region Y:<br>- ARI and SARI RSV testing data of base region: used for *Nyanza region*<br>- testing data from Siaya County Referral Hospital (CRH) and Kakamega CRH used for *Western Province*<br>- Kenyatta National hospital (NH), Nakuru CRH, Nyeri CRH used for *Rift valley, Central and Nairobi regions*<br>- national average including testing from Marsabit CRH and Coast General Teaching and Referral Hospital (GTRH) used for *Eastern and North Eastern regions* | $IF_{M,Y}=I_{M,Y}\times F_Y$<br><br>$IF_{NH,Y}=I_{NH,Y}\times F_Y$<br>**$F_Y$** = Proportion of ARI due to RSV in region Y<br><br>Nasopharyngeal and Oropharyngeal (NPOP) samples were systematically tested in Asembo for all enrolled cases in the study. This provided the RSV positivity for the base (Nyanza) region.<br>The ratio of RSV-positivity for ARI and SARI in the base region was applied to the SARI testing data of other regions to get the RSV-positivity for ARIs in the other regions.<br>The percent of ARIs positive for RSV |

| | | | | |
|---|---|---|---|---|
| | | | | in the base region was aggregated for the entire period with data (2010-14) and grouped into 3-month age bands from <3 months of age to 23 months, and 12-month age bands from 24 months of age. |
| **$I_B$** = Annual base rate of hospitalized SARI | SARI Definition: acute respiratory infection (onset within 10 days) with cough and reported or documented fever of ≥38C. For non-hospitalised severe cases, pneumonia was used as a proxy for SARI.<br><br>For non-hospitalised SARIs pneumonia was used as a proxy for healthcare seeking behavior. Pneumonia was defined as a cough and difficulty breathing for >2 days or a diagnosisa of pneumonia by a HCW (within the last year, when identified retrospectively). | KEMRI | Data collected from children <5 years visiting Kilifi CRH and residency data from participants in the Kilifi *Health and Demographic Surveillance System* (HDSS) [23]. | **$I_B = SARI_B/POP_B$**<br><br>$I_B$ was calculated for participants of the Kilifi HDSS for the years 2010-18 by dividing the age-specific number of hospitalized SARI by the age-specific population of HDSS residents in Kilifi.<br><br>$SARI_B$ = Total number of cases meeting SARI case definition hospitalized in surveillance catchment area<br><br>$Pop_B$=Population of surveillance catchment area |
| **$I_{H,Y}$** = Incidence of hospitalized SARI in region Y | SARI | KEMRI | Projecting the SARI rate of base region by using:<br>- regional data for healthcare seeking | $I_{H,Y} = I_B \times Adj_Y$<br>$Adj_Y = \left(1 + \sum_i (P_{i,Y} - P_{i,B}) \times (RR_i - 1)\right) \times \frac{DHS_Y}{DHS_B}$ |

| | | | | |
|---|---|---|---|---|
| | | | behavior from KHDS [21][22]<br>- regional data for risk factors | Risk factor prevalence $\mathbf{DHS_B}$ and $\mathbf{DHS_Y}$ from KHDS [21][22]. Risk factor contribution to SARI from [22].<br>Relative risk of HIV estimated 3.1% (95% CI, 1.2–8.1) from [18].<br>Credible intervals calculated from 1000 iterations allowing each risk factor to vary within a binomial distribution defined by the proportion with the characteristic and numbers interviewed. |
| $\mathbf{I_{NH,Y}}$ = Incidence of non-hospitalized SARI in region Y | SARI | KEMRI | From $\mathbf{I_{NH,Y}}$ by using data on healthcare seeking for acute respiratory illness from the 2018 Health Utilization Survey (HUS) data (unpublished) conducted in four counties (Siaya [Nyanza], Marsabit [Eastern], Nakuru [Rift Valley], Kakamega [Western]).<br><br>Proportion of individuals hospitalised with pneumonia was used as a proxy for SARI hospitalisation. Pneumonia definition used was: cough AND difficulty breathing >2 days OR a diagnosis by a HCW within the last year.<br>For Nyanza, Eastern, Rift Valley and Western regions the HUS estimates were used.<br>For Central, Nairobi and North Eastern the HUS estimates are adjusted by the care-seeking behavior of these | $I_{NH,Y}=\left(I_{H,Y}\times\frac{1}{HUS_Y}\right)-I_{H,Y}$<br>$\mathbf{HUS_Y}$ = proportion of all SARI cases hospitalised in region *Y*.<br>$HUS_Y=HUS_{2018}\times\frac{DHS_Y}{DHS_{2014}}$<br>$\mathbf{HUS_{2018}}$ = Proportion of all hospitalized SARI cases in the four counties<br>$\mathbf{DHS_Y}$ = Proportion of ARI cases seeking care in region Y (from KDHS).<br>$\mathbf{DHS_{2014}}$ = Proportion of ARI cases seeking care nationally (from KDHS)<br>Using the $\mathbf{DHS_Y}$/$\mathbf{DHS_{2014}}$ ratios assumes that healthcare seeking for SARIs varies by region proportionally to healthcare seeking in the case of ARIs. |

| | | | | |
|---|---|---|---|---|
| | | | regions compared to national average from KDHS [21][22]. | |
| Incidence of hospitalized/non-hospitalised RSV-associated SARI in region Y | SARI | KEMRI | Rates of hospitalised/ non-hospitalised SARI (**$I_{H,Y}$**, **$I_{NH,Y}$**) estimated as described above.<br><br>RSV positivity estimates:<br>- For Coastal and Nyanza regions: RSV positivity is from systematic testing of NPOP samples from hospitalised SARI patients.<br>For other regions testing was intermittent.<br>- Western region: Siaya CRH testing data was combined with Kakamega CRH.<br>- Rift Valley, Central and Nairobi regions: data from Kenyatta National hospital (NH), Nakuru CRH and Nyeri CRH were combined.<br>- Eastern and North Eastern regions: the national average was used combining the above data sources with Coast GTRH. | $IF_{H,Y} = I_{H,Y} \times F_Y$<br>**$F_Y$** = Proportion of SARIs testing positive for RSV<br><br>$IF_{NH,Y} = I_{NH,Y} \times F_{,Y}$ |
| **$I_{B\_SA,i}$** : Incidence of hospitalised SARI cases per population in base provinces, for age group *i* | SARI definition: children admitted to hospital with a physician's diagnosis of LRTI, with or without fever and a duration of 10 days or less. | National Institute For Communicable Diseases Of South Africa (NICD) | Two data sources described in [25]:<br>1) <u>Hospital surveillance</u>: RSV surveillance among patients with ILI, SARI and SARI in 3 public hospitals in 2 provinces, population of 457,000 (1.3% of South Africa) in 2015.<br>Sites are:<br>- Edendale Hospital (KwaZulu-Natal) | **$I_{B\_SA,i}$** = $[SARI_{enrolled} * (7/5) * (1/X_{SARI\_i}) * (1/HUS_{SARI})]/Pop_i$<br><br>**$SARI_{enrolled}$**: age-specific number of SARI cases enrolled<br>The factor 7/5 is used to adjust for non-enrolment over weekends. |

| | | | | |
|---|---|---|---|---|
| | | | - Klerksdorp and Tshepong Hospitals (North-West)<br><br>Respiratory specimens collected from all enrolled patients (ILI, SARI, SARI) and tested for 10 different viruses, among them RSV.<br><br>2) <u>Healthcare utilization surveys</u>: from studies of healthcare seeking behavior among individuals with reported ILI or SARI in three South African communities. | **$X_{SARI\_i}$**: proportion of all enrolled SARI cases belonging to age group *i*<br>$HUS_{SARI}$: proportion of SARI cases that sought care at the surveillance site out of SARI cases that sought care in any hospital obtained from the HUSs conducted in the catchment area of surveillance site [26], [27]. |
| **$I_{H,Y}$**: Incidence of hospitalised SARI cases per population in other provinces, for age group *i* | SARI | NICD | Risk factors of pneumonia for the other 7 provinces were taken from the DHS [22,23].<br>These included HIV infection, exposure to indoor air pollution, crowding, malnutrition, low birth-weight and non-exclusive breastfeeding (last 3 only for children <5yr). SARI-association of risks from [18], [28], [29].<br><br>Provincial rates were also adjusted by proportion of ARI cases seeking care in the given province relative to the base provinces [30], [43], ie. healthcare-seeking among ARIs was used as a proxy for SARIs. | **$I_{H,Y}$** $= I_{H,B} * Adj_Y * DHS_{H,Y}/DHS_{H,B}$<br><br>**$Adj_Y$**$=1+\sum_{i=1}^{n}(P_{i,Y}-P_{i,B})(RR_i-1)$<br>**$DHS_{H,Y}$**: proportion of ARI cases seeking care in province Y (from DHS [18], [28], [29])<br>**$DHS_{H,B}$**: proportion of ARI cases seeking care in the base provinces |
| **$IF_{H,Y\_SA}$**: RSV-associated SARI hospitalisation in all provinces | SARI | NICD | Testing procedure described in [19]. Respiratory specimens were collected from all enrolled patients, and were tested for the presence of 10 | **$IF_{H,Y\_SA}$**$= I_{H,Y}*F_Y*F_{Y,AF}$<br><br>**$I_{H,Y}$**: SARI hospitalisation rate in province Y (including the base |

| | | | | |
|---|---|---|---|---|
| | | | respiratory viruses (including RSV) using a multiplex real-time reverse-transcription PCR assay. | provinces)<br>**$F_Y$**: proportion of SARI cases testing positive for RSV [26]<br>**$F_{Y,AF}$**: attributable fraction of RSV virus detection to illness, obtained at the same sentinel sites [44] |
| **$I_{NH,B\_SA}$**: Incidence of non-medically-attended SARI cases per population in base provinces | SARI | NICD | From HUS data [26], [27] | **$I_{NH,B\_SA}$**$=I_{H,B\_SA}*(1/HUS_B - 1)$<br><br>**$HUS_B$**: proportion of all SARI cases that are hospitalized in the base provinces |
| **$I_{NH,Y\_SA}$** : Incidence of non-medically-attended SARI cases per population in other provinces | SARI | NICD | From HUS data [26], [27] and estimates on risk factors | **$I_{NH,Y\_SA}$**$=I_{NH,B\_SA}*Adj_Y * DHS_{NH,Y}/DHS_{NH,B}$<br><br>(all variables already defined above) |
| **$IF_{H,Y\_SA}$**: RSV-associated non-hospitalised SARI rate in all provinces | SARI | NICD | From previous variables and RSV testing data at sentinel sites | **$IF_{NH,Y\_SA}$**$= I_{NH,Y}*F_Y*F_{Y,AF}$<br><br>**$I_{NH,Y}$**: SARI hospitalisation rate in province Y (including the base provinces)<br>**$F_Y$**: proportion of SARI cases testing positive for RSV<br>**$F_{Y,AF}$**: attributable fraction of RSV virus detection to illness, obtained at the same sentinel sites [45] |
| **$ILI_i$**: age-specific rate of | Definition: outpatient of any age presenting with | NICD | From SARI rates, combined with proportion of ILI consultations | **$ILI_i$** $= SARI_i * Z * X * (Y_{ILI\_i} / Y_{SARI\_i})$ |

| | | | | |
|---|---|---|---|---|
| RSV-associated ILI outpatient consultations in age group *i* | either temperature ≥38C or history of fever and cough of a duration ≤10 days.<br>We approximated ARIs from ILIs by taking the proportion of ARIs with fever (on average 23.3% from 2015 to 2017) in Kenya [32] and dividing the ILI rates by this proportion. | | referred to hospital and proportion of SARI cases that previously seeked outpatient care | $\mathbf{SARI_i}$: age specific rate of SARI hospitalization (adjusted for non-enrollment and healthcare seeking behavior)<br>**Z**: proportion of SARI cases that sought outpatient care before hospitalization<br>**X**: ratio of ILI consultation referred to hospital to the total number of ILI consultations obtained from ILI surveillance over the study period<br>$\mathbf{Y_{ILI\_i}}$: proportion of ILI cases in age group ***i*** over the total number of ILI cases after adjusting for non-enrolment<br>$\mathbf{Y_{SARI\_i}}$: proportion of SARI cases in age group ***i*** over the total number of SARI cases after adjusting for non-enrolment |

**SI Table 2**: Update parameter values used in the analysis

| Parameter | Type | value | Source |
|---|---|---|---|
| **Efficacy** | Maternal vaccination, using Novavax clinical trial data | Symptomatic RSV LRTI: 39.4% (CI95: 5.3%, 61.2%)<br>RSV LRTI with hospitalisation: 44.4% (CI95: 19.6%, 61.5%)<br>RSV LRTI with severe hypoxemia: 48.3% (CI95: -8.2%, 75.3%) | [6] |
| **Efficacy** | Monoclonal antibody | Symptomatic RSV LRTI: 70.1% (CI95: 52.3%, 81.2%)<br>RSV LRTI with hospitalisation: 78.4% (CI95: 51.9% to 90.3%) | [12] |
| **Duration of protection** | Maternal vaccination | Default case: 3 months of protection with efficacy figures above.<br>With exponential waning vaccine efficacy is a function of time; parameters in SI Table 3. | [6] |
| **Duration of protection** | Monoclonal antibody | Default case: 5 months protection with efficacy figures above, 0% protection after.<br>With exponential waning vaccine efficacy is a function of time; parameters in SI Table 3.<br>See Supplementary Methods for calculation. | [12] |
| **Cost** | Hospitalisation, South Africa | See SI Figure 1, between 559 and 1032 USD per illness episode | own data |
| **Cost** | Outpatient care, South Africa | 25 USD (CI95: 18.3, 31.8) | own data |
| **Cost** | Outpatient care, Kenya (Siaya) | 20.91 USD (calculated from total cost to the hospital at Siaya, total number of inpatient and outpatient cases, and an assumed ratio of inpatient to outpatient costs of 4.9) | own data |
| **Cost** | Inpatient care, Kenya | 102.46 USD (see above) | own data |
| **Cost** | Costs | mean: 34.67USD (median: 14.23, CI95 | own |

| | | | |
|---|---|---|---|
| | incurred to households prior to hospitalisation, Kenya (Siaya) | 12.86-56.49) | data |
| **Cost** | Costs incurred to households during hospitalisation, Kenya (Siaya) | Mean: 138.51USD (median: 122.7, CI95: 106.54-170.48) | own data |
| **Cost** | Costs incurred after hospitalisation, Kenya (Siaya) | 15.35USD (median: 4.61, CI95: 0-34.46) | own data |
| **Cost** | Number of inpatients in 12 months, Kenya (Siaya) | 7330 | own data |
| **Cost** | Number of outpatients in 12 months, Kenya (Siaya) | 62604 | own data |
| **Cost** | Total healthcare cost in 12 months | 2,060,208 USD | own data |
| **Cost** | Maternal vaccination | 3, 10, 30 USD per dose | own data |
| **Cost** | Monoclonal antibody | 6, 20, 60 USD per dose | own data |

**SI Table 3**: parameter estimates for exponentially waning efficacy (fits to efficacy figures in SI Table 2, sources are the same)

| Intervention | Endpoint | *c* | *α* (in months) |
|---|---|---|---|
| MV | symptomatic RSV LRTI | 0.627 | 0.57 |
| MV | RSV LRTI with hospitalisation | 0.706 | 0.57 |
| MV | RSV LRTI with severe hypoxemia | 1 | 1.1 |
| mAb | Symptomatic RSV LRTI | 1 | 0.196 |
| mAb | RSV LRTI with hospitalisation | 1 | 0.131 |

**SI Table 4**: Parameter estimates for fits of efficacy data with beta distributions. Fits were performed by the *optim* function of R. The numerical values of fitting parameters will vary with individual fits (fits to efficacy figures in SI Table 2, sources are the same)

| **Drug** | **Disease type** | **Mean (CI95)** | **Beta(α,β)** (if the distribution is shifted: **shift**, **scale**) |
|---|---|---|---|
| Maternal vaccine candidate from Novavax [7] | Symptomatic RSV LRTI | 39.4% (5.3, 61.2%) | 7.586, 11.667 |
| Maternal vaccine candidate from Novavax | RSV LRTI with hospitalisation | 44.4% (19.6, 61.5%) | 12.023, 15.055 |
| Maternal vaccine candidate from Novavax | RSV LRTI with severe hypoxemia | 48.3% (-8.2, 75.3%) | 36.845, 39.439 (-1.473, 3.742) |
| Monoclonal antibody Nirsevimab, published interim results [12] | Symptomatic RSV LRTI | 70.1% (52.3, 81.2%) | 21.38, 9.119 |
| Monoclonal antibody Nirsevimab, published interim results [12] | RSV LRTI hospitalisation | 78.4% (51.9, 90.3%) | 9.333, 2.571 |
| Maternal vaccine candidate from Pfizer, unpublished interim results [13] | Medically attended LRTI | 84.7% (21.6, 97.6%) | 1.445, 0.261 |
| Maternal vaccine candidate from Pfizer, unpublished interim results | Medically attended LRTI | 91.5% (-5.6, 99.8%) | 0.19, 0.018 (shift=-0.057, scale=1.057) |

### Supplementary Methods

***Comparison with parameter estimates in Li et al***

Hospitalisation probabilities in Li et al. [15] were assumed to be identical across age groups (mean value of 8.7%) based on Nokes et al. [46], resulting in an identical age distribution for all hospitalised cases, with uncertainty accounted for by doubling the standard deviation.

Using generalised additive mixed models, the Li et al. analysis inferred an age-specific incidence curve of RSV disease from the data in the global RSV meta-analysis by Shi et al [3]. The incidence curve was calibrated to peak at 6 months of age at a value depending on the country-specific total incidence, which was in the range of 100-150 disease episodes per 1000 person-years. Mean duration of illness and hospital stays were sampled from gamma distributions (means: 11.2 and 5.8 days). Efficacy, duration of protection and cost estimates for MV and mAb are shown in SI Table 2.

We updated these estimates with our hospital-based incidence data, which included estimates for the probability of hospitalisations and in the case of Kenya also RSV-associated deaths per population. The main difference is in the age distribution of SARI cases being markedly different in our dataset from the age distribution of non-severe (ARI) cases, which changes the rate of hospitalisations and deaths.

***Fitting confidence intervals of efficacy estimates by beta distributions***

We perform a probabilistic sensitivity analysis to account for uncertainties in parameters, including the efficacy of the preventive biologics used. To obtain a probability distribution we can sample from, we use a beta distribution to fit the CI95 values of the efficacy figures reported from clinical trials (SI Table 2, 4). We used the *optim* function in R to find the beta distribution's two parameters (α,β), sampling through a range of initial values of α and β that satisfy the constraint for the mean efficacy, $E(X)=\alpha/(\alpha+\beta)$.

In some cases the lower bound of the confidence interval of the efficacy estimate has a negative tail, which cannot be fit by a beta distribution. In these cases we introduced two other fitting parameters, a 'scaling parameter' and a 'shifting parameter', so that we are fitting an adjusted beta distribution:

*rbeta(α,β)*scaling_parameter + shifting_parameter*

The fits are in SI Table 4. In the case of the efficacy figure 'RSV LRTI with severe hypoxemia' for MV, the mean and the confidence interval cannot be well fit simultaneously. Since the fitting is performed using the confidence intervals, the estimate for the mean is lower than the mean in the data (35% and 48%, respectively).

***Exponential waning model for efficacy figures of MV and mAb***

We fit MV and mAb efficacy figures with an exponential decay model by setting two constraints. First, the half-life ($t_{0.5}$) was fixed to the half-life of antibodies for MV and to mAb serum concentration observed in the clinical trials [6], [12]. For MV the half-life of antibodies is 36.5 days (Table S12 in [6]), whereas for mAb serum concentration it is 59.3 days. The second constraint is that the exponential curves need to have a mean value corresponding to the efficacy figures in the 0-90 days (MV) and 0-150 days (mAb) intervals.
The period of protection is denoted as $t_{dur}$ and was set to 90 days in the case of MV and 150 for mAb.
There are two parameters to be determined, one is a scaling constant (c) for the efficacy and the other is the rate of decay (α). The two parameters are then constrained as:

$$\exp(-\alpha t_{0.5}) = 0.5 \qquad \text{(I)}$$

$$\frac{1}{t_{dur}} c \int_0^{t_{dur}} \exp(-\alpha t)\, dt = \text{VE} \qquad \text{(II)}$$

Reordering, we have for the two parameters:

$$\alpha = \log(2)/t_{0.5}$$

$$c = \alpha \frac{t_{dur} VE}{1 - \exp(-\alpha t_{dur})}$$

In some cases this calculation yields a *c* value above 1 (which would mean a more than 100% efficacy). In this case we fixed *c* to 1, and calculated *α* as

$$1 = \alpha \frac{t_{dur} VE}{1 - \exp(-\alpha t_{dur})}$$

yielding

$$\alpha t_{dur} + \log(\alpha) = \frac{1}{t_{dur} VE}$$

Which we solved numerically for *α.* This calculation leads to half-lives slightly above the values reported in the clinical trials, but the same mean efficacy value over the relevant period.
The values of *c* and *α* for MV and mAb with the different endpoints are shown in SI Table 3.
